# Pallidal Local Field Potentials markers of dystonia aetiology and severity in children

**DOI:** 10.64898/2026.10.03.26364638

**Authors:** Irene Faiman, Adela Desowska, Sarah Perides, Margaret Kaminska, Claire Murray, Sinead Barkey, Claire Thompson, Katy Wood, Aoife Flynn, Keyoumars Ashkan, Harutomo Hasegawa, Daniel E. Lumsden, Jean-Pierre Lin, Verity McClelland

## Abstract

**Background:** Aberrant basal ganglia activity is a key feature of the pathophysiology of dystonia, but evidence is predominantly based on adult cohorts. Neurophysiological signatures during development remain poorly characterised. Local field potentials (LFPs) recorded by bidirectional neurostimulators provide a direct measure of globus pallidus internus (GPi) activity. In a cross-sectional paediatric cohort, we investigated whether pallidal oscillatory, aperiodic and connectivity characteristics are associated with dystonia severity and whether these differ between genetic and acquired dystonias.

**Methods:** We recruited 30 children and young adults (5–20 years) with medically refractory dystonia (genetic *n* = 10; acquired *n* = 20) who underwent bilateral GPi Deep Brain Stimulation (DBS) implantation. Ten minutes of pallidal LFPs were recorded using Medtronic Percept™ RC neurostimulators with stimulation off, before DBS activation and, where available, at 12 months. Spectral power, aperiodic exponent and interhemispheric pallidopallidal coherence were quantified across delta (1–4 Hz), theta (4–8 Hz), alpha (8–13 Hz), beta (13–30 Hz), and low-gamma (30–45 Hz) frequency bands. Linear mixed-effects models were implemented to establish whether LFP markers were predictive of dystonia severity, as measured by the Burke–Fahn–Marsden Dystonia Rating Scale (BFMDRS-Movement). Firth’s Bias-Reduced Logistic Regression assessed whether LFP markers were predictive of dystonia aetiology.

**Results:** Spectral peaks were most consistently detected in the delta, beta, and low-gamma bands. Delta and low-gamma peak power significantly predicted dystonia severity. This association was independent of medication burden, structural GPi involvement, and dystonia aetiology. Theta and alpha peaks were detected less consistently, and their power was not associated with dystonia severity. Pallidopallidal coherence significantly predicted dystonia severity in interaction with medication load in all frequency bands. Low-gamma peak power was significantly lower in acquired than genetic dystonia, but the potentially confounding influence of dystonia severity could not be excluded. The aperiodic exponent was not associated with severity or aetiology.

**Conclusion:** Pallidal LFP markers are associated with dystonia severity in children, with delta peak power, low-gamma peak power and pallidopallidal coherence reflecting clinically relevant features of disease expression. These findings highlight the potential of pallidal LFPs to characterise dystonia across the lifespan and motivate longitudinal studies of their developmental and clinical trajectories.

## Introduction

Dystonia is movement disorder characterised by sustained or intermittent abnormal movements, postures, or both^1^ often leading to severe motor disability. It can arise from diverse genetic or acquired causes,^1^ with dystonic/dyskinetic cerebral palsy (CP) representing a frequent cause of acquired dystonia in childhood.^2^ Although current models implicate dysfunction within a distributed network including the basal ganglia, cerebellum, thalamus and sensorimotor cortex, the pathophysiology of dystonia is not fully understood.^3–5^ Bilateral deep brain stimulation (DBS) of the globus pallidus internus (GPi) is an established treatment for medically refractory dystonia, but clinical outcomes vary considerably, with people with genetic dystonias generally demonstrating greater benefit than those with acquired aetiology.^6–8^ The pathophysiological basis for this heterogeneity in DBS response remains unclear.^6^

Local field potential (LFP) recordings obtained from temporarily externalised DBS leads in the peri-operative period and, more recently, from bidirectional neurostimulators provide a direct measure of the electrophysiological activity of subcortical structures.^9^ Previous studies have described abnormalities across a range of oscillatory frequencies in GPi LFPs recorded from people with dystonia.^10,11^ Among these, low-frequency (4–10 Hz) power spectral peaks^10^ and low-frequency coherence between different nodes of the dystonic network^11^ have been reported most consistently. Multiple studies have also demonstrated associations between low-frequency GPi activity and dystonia severity, including correlations with clinical rating scales,^12–14^ involuntary muscle activity measured by electromyography,^15–18^ and symptom fluctuations.^19,20^ These observations, together with evidence of exaggerated low-frequency drive to muscles,^21–23^ suggest that low-frequency oscillations may represent an important feature of the pathophysiology of dystonia, with the hypothesis that DBS might overwrite or regularise these pathological activity patterns.^18^

Several studies have reported similar low-frequency GPi activity across different dystonia aetiologies such as genetic and acquired,^10^ and across groups with different dystonia distributions including generalised, segmental and focal.^11^ Additionally, similar findings in patients with other hyperkinetic presentations, such as Parkinson’s Disease with levodopa-induced dyskinesia^24^ and Tourette’s syndrome^25^ have led to the suggestion that excessive low frequency power might not be disease-specific, but rather reflective of hyperkinetic symptomatology.^5^ These observations suggest that GPi oscillations may primarily reflect the expression and severity of dystonic symptoms rather than the underlying mechanism of disease.

However, evidence supporting this interpretation is limited by small sample sizes, a predominance of adults with idiopathic or rare genetic dystonias in the LFP literature, and a lack of direct comparisons across aetiologies.^10,11^ This interpretation also remains difficult to reconcile with the observed differences in DBS responsiveness between genetic and acquired dystonias,^6^ highlighting the need to ascertain whether GPi LFP patterns represent shared electrophysiological signatures across aetiologies. Clarifying this relationship would be critical to informing aetiology-specific treatment strategies with potential for improved clinical outcomes.

This question is particularly important in the context of the developing brain, where early brain injury and maturation shape the organisation and plasticity of the network underlying dystonia, and where activity in critical neurodevelopmental windows has enduring effects which may only manifest later in life. However, the paediatric DBS literature has been limited to small retrospective cohorts,^6,7,26^ with larger studies just recently emerging.^8^ Paediatric LFP studies are also extremely limited so far, with only a small number of reports to date, each including no more than ten patients.^19,20,27–29^ Defining whether electrophysiological signatures identified in adult cohorts apply to the developing brain and across dystonia aetiologies is essential to understand the generalisability of the proposed physiomarkers and their potential clinical application across the lifespan.

This study examines GPi LFP recordings from a single-centre paediatric dystonia cohort, including a substantial group with dystonic CP. Our objectives were to assess i) whether the association between low-frequency LFP activity and symptom severity in adults can be replicated in children and young people with dystonia, and ii) whether different aetiologies exhibit distinct electrophysiological signatures despite a shared clinical phenotype.

## Materials and methods

### Participants and surgical procedure

We recruited a consecutive sample of 30 children and young adults (age between five and 20; *n* = 3 over 18 years) who underwent bilateral pallidal DBS implantation (Medtronic Percept^TM^ RC neurostimulator) at Evelina London Children’s Hospital (ELCH), Guy’s and St Thomas’ NHS Foundation Trust (Jan 2024–Jan 2026) for management of dystonia symptoms. Suitability for DBS was determined following multi-disciplinary clinical assessment at ELCH; the decision to implant followed established clinical pathways, as previously reported.^30^ Implantation was performed under general anaesthesia using direct MRI-guided targeting and the Neuromate (Renishaw, UK) robot. Lead placement was confirmed with the O-arm intra-operative imaging system. The Implantable Pulse Generator (IPG) was buried in a subcutaneous pocket at right abdominal (*n* = 27) or chest level (*n* = 3).

Ethical approval was obtained to use Percept BrainSense data for research (East Midlands - Nottingham 1 Research Ethics Committee; REC: 23/EM/0035, IRAS ID: 317454, latest amendment: 03/02/2025). All participants or their parent/guardian gave written informed consent.

### Clinical data collection

Demographic and clinical data were extracted from clinical notes, including the Burke– Fahn–Marsden Dystonia Rating Scale (BFMDRS)^31^ –Movement and Disability scores, which assess dystonia severity and associated disability at baseline (pre-DBS) and 12-months post-DBS; the five-level Gross Motor Function Classification System (GMFCS)^32^ and the Manual Ability Classification System (MACS),^33^ assessing mobility/gross motor skills, and object handling, respectively; the Canadian Occupational Performance Measure (COPM)^34^ –Performance and Satisfaction scales, capturing patients’ and/or caregivers’ perspectives on self-care, productivity and leisure (when two raters were present, the average was used); the Caregiver Priorities and Child Health Index of Life with Disabilities (CPCHILD),^35^ measuring caregivers’ perspectives on health-related quality of life. In addition, the Clinical Global Impression –Severity (CGI-S)^36^ scale rated dystonia severity from one to seven (Supplementary Section 1). CGI ratings were independently completed at each time point by the same consultant paediatric neurologist and specialist nurse, with the mean score reported.

Adapted from McClelland *et al.*,^37^ brain MRI findings were categorised from pre-DBS neuroradiology reports as (1) normal, (2) evidence of white matter injury (including periventricular leukomalacia, ventricular enlargement, white matter signal change, reduced white matter volume), (3) term Hypoxic Ischaemic Encephalopathy (HIE), (4) know metabolic pattern associated with clinically diagnosed metabolic disorders, (5) focal infarct, (6) malformations of cortical development (e.g., focal cortical dysplasia), and (7) other structural abnormalities.

Information on the presence and normality of motor evoked potentials (MEPs), central motor conduction times (CMCT), and somatosensory evoked potentials (SEPs) was extracted from neurophysiology reports.

### LFP data acquisition

Pallidal LFP recordings were acquired as part of routine clinical practice prior to DBS activation (either three to five days, *n* = 25, or one-month postoperatively, *n* = 5), with additional recordings at 12 months post-implantation in 14 participants who completed clinical follow-up. Recordings comprised an average of 10.0 min (SD: 4.2; range: 2.1– 13.5) of Indefinite Streaming data acquired simultaneously from both hemispheres (four level-wise contacts per lead, excluding segments, giving six bipolar recordings per hemisphere; Fig. 1) during spontaneous activity. Participants were seated or lying and awake during recordings – either resting or during spontaneous activity – except for two recordings obtained during sleep. Stimulation is automatically turned off during Indefinite Streaming. During recordings, BrainSense automatically applies two low pass filters (100Hz), and two high pass filters (1Hz). No additional offline filtering was applied.

**Figure 1.**
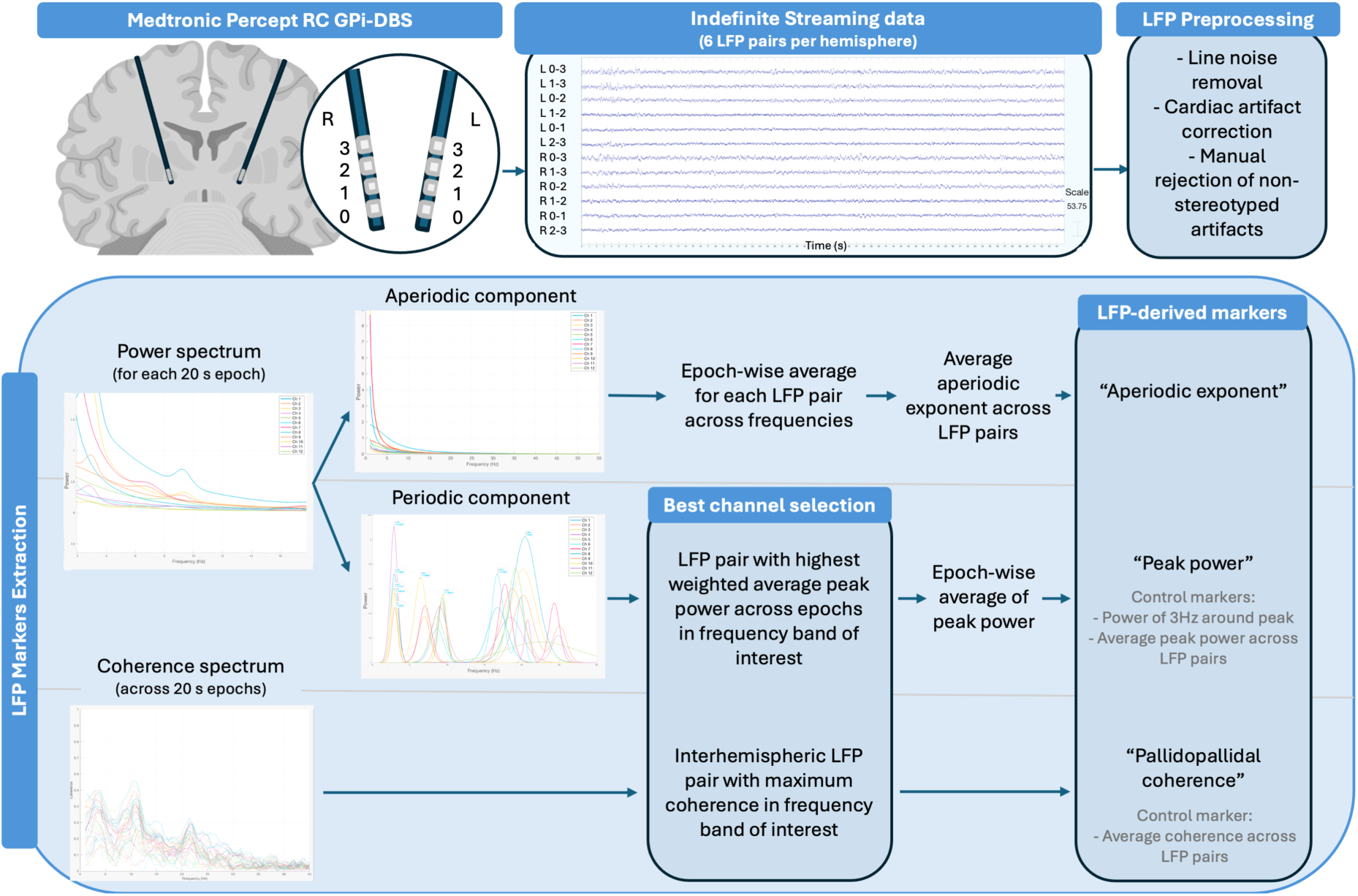
Visual summary of LFP acquisition and analysis pipeline. Indefinite streaming data are acquired as bipolar recordings with stimulation off, with two contacts within the same electrode serving as the anode and cathode. Each lead comprises eight physical contacts arranged in four levels; the segmented contacts within each level are treated collectively as a single “level” contact (0-3), resulting in four level contacts per hemisphere for analysis. All possible level-wise combinations are simultaneously recorded resulting in six bipolar LFP channels per hemisphere. LFP data are preprocessed, and markers are derived to be tested as predictors of dystonia severity or aetiology in subsequent statistical analyses. The raw data and spectra shown for illustrative purposes are from a single participant.

### Offline LFP preprocessing

Line noise at 50 and 100 Hz was removed using the cleanLineNoise MATLAB function, with 4s taper windows (1s slide), 2Hz search bandwidth, and default iterative and statistical settings.^38^ If present, cardiac artifacts were corrected using a custom template subtraction pipeline based on Hammer *et al.*^39^ In-detail description of this method is provided in Supplementary Material; the resulting MATLAB function is publicly available at [link to repository will be included once the manuscript is accepted]. Cardiac artifacts were infrequent, detected and subsequently removed from 26 LFP channels from seven Indefinite Streaming recordings only. Data were then visually inspected and non-stereotyped artifacts manually rejected.

### LFP-derived markers

#### Peak power and aperiodic exponent

Preprocessed LFP data were segmented into 20 s continuous epochs, and the *specparam* algorithm was applied via the MATLAB function eeg_htpCalcFooof (vhtp toolbox).^40^ *Specparam* models aperiodic 1/f components and periodic gaussians (oscillatory peaks) for each epoch’s power spectrum, previously computed between 1 and 45 Hz using a Short-Time Fourier Transform (spectrogram) with 1-s Hamming window and 50% overlap, and time-wise averaging of the time-resolved spectra. Parameters included peak width between 0.5 and 12 Hz, maximum number of peaks equal to five, peak threshold of 2 SD, and modelling the aperiodic knee, i.e., the point marking a change in the aperiodic spectral slope. The “best LFP channel” was then selected as follows. For each LFP channel, *specparam*-modelled linearised peaks in the theta band (4–8 Hz) were extracted (with only the highest peak considered in case of multiple peaks), and a weighted average (*x̂*) of peak power across epochs for each channel *i* was calculated as

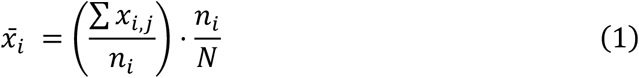

where *x_i,j_* is the peak power in epoch *j* for channel *i, n_i_* is the number of epochs where a peak was detected, and *N* is the total number of epochs. This operation scales each channel’s peak power mean by the proportion of epochs in which a peak was detected, the latter serving as a “peak reliability factor” that reduces the influence of channels with infrequent theta peaks. As a result, channel selection reflects a balanced consideration of both magnitude and prevalence of theta peaks. The LFP channel with the highest weighted average was selected for further processing (Fig. 1). A small-sample-bias-adjusted^41^ coefficient of variation was calculated to quantify variability in weighted average between LFP channels.

For the selected channel, average peak power across epochs at the *specparam*-identified peak frequency was extracted from the original power spectrum following subtraction of the aperiodic component (including the knee).

Analyses were repeated using peak frequency search windows in the delta (1–4 Hz), alpha (8–13 Hz), beta (13–30 Hz), and low-gamma (30–45 Hz) ranges.

The epoch-wise average *specparam*-modelled aperiodic exponent was computed and averaged across channels (Fig. 1); since the model included a knee, the exponent is a measure of the aperiodic component past the knee frequency; the median (IQR) knee frequency is reported.

#### Pallidopallidal coherence

As homotopic interhemispheric connectivity occurs with zero-or near-zero phase delay, a zero-phase inclusive method was selected.^42,43^ Pallidopallidal coherence (*C*_xy_), which quantifies how consistently two LFP signals *x* and *y* maintain a linear relationship (in both amplitude and phase) at a given frequency *f*, was therefore calculated as:

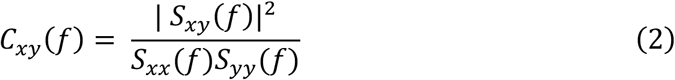

where S_xy_ represents the cross-spectrum, S_yy_ and S_xx_ the auto-spectra. This was computed using Fieldtrip ft_connectivityanalysis from complex Fourier coefficients obtained via multitaper spectral analysis (all available 20 s segments split into non-overlapping 2 s epochs and analysed using the mtmfft method with DPSS (Slepian) tapers and 2Hz frequency smoothing window), with cross-spectral and auto-spectral estimates averaged across epochs.

As subtle variations between the positioning of left and right GPi electrodes may occur, all interhemispheric combinations were extracted, and the LFP pair displaying maximum coherence in each frequency band was used for subsequent analyses (Fig. 1), consistent with previous work.^12^

### Statistical analyses

To test whether LFP-derived measures predicted dystonia severity, linear mixed-effects models^44^ with Restricted Maximum Likelihood estimator were implemented. These allow increasing power by leveraging repeated measures (*n* = 14 with 12-months assessment), handling missing follow-up data (*n* = 16), and modelling between-patient variability as a random intercept. Multiple models were fitted with theta peak power, aperiodic exponent, or theta pallidopallidal coherence as fixed effects and BFMDRS– Movement score as predicted variable, with a random intercept for participants. Confidence intervals and *P*-values for fixed-effects were obtained using parametric bootstrapping (5,000 iterations). For natural spline terms, statistical inference was based on likelihood ratio tests^45^ comparing models with and without the spline term, as individual spline-basis-functions coefficients are not independently interpretable. Competing models were compared using likelihood ratio tests^45^ assessing improvement in model fit. Model assumptions were evaluated using simulation-based residual diagnostics (‘DHARMa’ package^46^ V0.4.7, 2024-10-16) and Levene’s test for homogeneity of variance.^47^

To test whether LFP-derived measures predicted dystonia aetiology, Firth’s Bias-Reduced Logistic Regression with penalised log-likelihood estimator was implemented.^48^ The Firth’s method produces nearly unbiased maximum likelihood estimates in small samples (*n* = 30), with unbalanced outcome classes (genetic dystonia *n* = 10; acquired *n* = 20) and low number of observations in cross-tabulation with the outcome (*n* < 5 for one covariate).^49^ As aetiology is stable over time, LFP-derived measures were averaged across timepoints (for *n* = 14 with repeated measures) to improve the stability of trait-level markers. The resulting theta peak power, aperiodic exponent, and theta pallidopallidal coherence estimates were entered as predictors in separate models, with aetiology as the predicted variable. Genetic dystonia was the reference category. Odds Ratios (ORs) and Confidence Intervals were derived to estimate the magnitude of the association.

With *n* = 30 participants (or *n* = 44 observations in mixed-effect models), statistical considerations allow a limited number of predictors (three to four).^50^ For all models, an a priori decision was made to include covariates with the highest potential to confound LFP estimates, namely number of psychoactive medications and pallidal involvement on structural MRI. Unadjusted analyses quantifying the association between LFP-derived markers and BFMDRS-M scores without any model covariates are reported. Whilst our primary hypothesis was focused on the theta frequency band, all analyses were repeated using peak power and pallidopallidal coherence extracted from delta, alpha, beta, and low-gamma; for each predictor type, False Discovery Rate (FDR) correction^51^ was applied across bands.

Data were analysed using R statistical software V4.5.3 (2026-03-11) with the ‘logistf’ package^52^ V1.26.1 (2025-04-16) and the ‘lme4’ package^53^ V.2.0.1 (2026-03-05). If present, the impact of covariates’ heteroscedasticity was assessed through a sensitivity analysis using a Gaussian model with a dispersion term allowing residual variance to vary by the covariate’s levels (‘glmmTMB’ package V 1.1.14, 2026-01-15).^45^

### Control analyses

Control analyses included computing mean power in a 3Hz narrow band surrounding the peak,^12^ and computing average peak power and pallidopallidal coherence across all LFP channels to exclude potential channel selection bias (Fig. 1).

To control for potential confounding between the two outcome variables, significant mixed-effects models predicting dystonia severity were re-estimated with aetiology as an additional covariate. Conversely, logistic regression models predicting dystonia aetiology were re-estimated with dystonia severity as an additional covariate.

For significant models, sensitivity analyses were run excluding two recordings acquired during sleep and two (from the same participant) with detected epileptiform discharges. Note that as epileptiform abnormalities were removed during preprocessing, none were present in any of the analysed data.

### Population descriptive statistics

Descriptive statistics are reported for the whole group and stratifying by aetiology. Normality was assessed using the Shapiro-Wilk test.^54^ For continuous normally distributed variables, Welch’s *t*-test^55^ estimated the significance of differences between aetiology groups; Mann-Whitney U test^56^ assessed other distributions. For categorical variables, Fisher’s exact test^57^ was used to account for low cell count (n < 5).

## Results

### Population description

Thirty-three families were invited and 32 consented to participate. Two were excluded due to unavailable Indefinite Streaming data, resulting in a final sample of 30 (Table 1).

**Table 1.** Demographic, clinical and LFP characteristics for the included population, with statistical group comparison between the subgroups with genetic and acquired aetiologies.

|  | Whole sample<br>*n* = 30 | Aetiology:<br>Genetic<br>*n* = 10 | Aetiology:<br>Acquired<br>*n* = 20 | Statistic | *P* value |
| --- | --- | --- | --- | --- | --- |
| **Demographic** |  |  |  |  |  |
| Male sex, *n* (%) | 13 (43.33) | 4 (40) | 9 (45) | Fisher's exact | 1.000 |
| Age at surgery, mean (SD) | 11.60 (3.73) | 12.62 (4.26) | 11.10 (3.44) | *t*(15.05) = 0.98 | 0.341 |
| Ethnicity, White (vs Asian and Black), *n* (%) | 16 (53.33) | 5 (50) | 11 (55) | Fisher's exact | 0.102 |
| Handedness, right (vs left and impossible to determine) *n* (%) | 15 (50.00) | 8 (80) | 7 (35) | Fisher's exact | 0.048* |
| Years in education, mean (SD) | 6.70 (3.57) | 8.00 (4.37) | 6.05 (3.02) | *t*(13.43) = 1.26 | 0.226 |
| School type, Special *n* (%) | 17 (56.67) | 2 (20) | 15 (75) | Fisher's exact | 0.007** |
| **Clinical** |  |  |  |  |  |
| Age at onset, median (IQR) | 1.00 (1.88) | 4.00 (4.38) | 0.00 (1.00) | *W* = 196.0 | <0.001*** |
| Disorder duration at surgery, mean (SD) | 9.52 (3.97) | 7.62 (4.36) | 10.52 (3.46) | *t*(15.12) = -1.82 | 0.088 |
| Temporal pattern of disorder, Static (vs Progressive) *n* (%) | 20 (66.67) | 1 (10) | 20 (100) | Fisher's exact | <0.001*** |
| Isolated dystonia (vs combined with other movement disorders) *n* (%) | 22 (73.33) | 7 (70) | 15 (75) | Fisher's exact | 1.000 |
| Pallidal involvement on MRI, n (%) | 17 (56.67) | 3 (30) | 14 (70) | Fisher's exact | 0.056 |
| Number of medications, median (IQR) | 3.00 (3.75) | 1.00 (1.75) | 4.00 (2.00) | W = 24.5 | <0.001*** |
| Baseline BFMDRS-Movement score, median (IQR) | 99.00 (35.00) | 62.00 (34.50) | 105.50 (15.50) | W = 28.0 | 0.003** |
| **LFP data** |  |  |  |  |  |
| Number of 20 sec segments analysed per person, median (IQR) | 26.50 (9.25) | 25.00 (11.50) | 28.00 (9.00) | W = 152.0 | 0.106 |
| Completed 12-months follow-up, n (%) | 14 (46.67) | 5 (50) | 9 (45) | Fisher's exact | 1.0000 |
| BFMDRS, Burke-Fahn-Marsden Dystonia Rating Scale. * $P < 0.05$ ** $P < 0.01$ *** $P < 0.001$ | | | | | |

#### Dystonia characteristics

Participants with genetic dystonia (*n* = 10) included autosomal dominant (DYT1/*TOR1A*, DYT6/*THAP1*, DYT11/*SGCE*, *KMT2B*, *EIF2AK2*), autosomal recessive (Glutaric aciduria type 1, GA-I/*GCDH*, HARP/*PANK2*), and mitochondrial (Leigh syndrome) aetiologies. The acquired subgroup (*n* = 20) consisted of people with dystonic/dyskinetic Cerebral Palsy due to prematurity (*n =* 13), kernicterus (*n* = 6; of which *n* = 3 secondary to underlying metabolic diagnoses), or term HIE (*n* = 1). For all participants, the clinical phenotype was characterised by persistent, generalised dystonia with bilateral involvement; three showed predominantly asymmetric distribution. The disorder was progressive in almost all participants with genetic dystonia and static in all with acquired dystonia (Table 1). Age at onset differed between genetic and acquired aetiologies, whereas dystonia duration at surgery was comparable between groups (Table 1). 73% of participants had isolated dystonia, with proportions consistent across aetiology groups (Table 1); concurrent motor features in the remainder included chorea (*n* = 3), athetosis (*n* = 4), spasticity (*n* = 1) and myoclonus (*n* = 3). Non-motor neurological comorbidities are in Supplementary Material Section 2.

#### Demographic characteristics

Demographic characteristics were comparable between genetic and acquired groups, except the acquired group included more people with undetermined handedness and more people attending special schools (Table 1).

#### Pre-DBS investigation profiles

MRI abnormalities were present in all but two participants, with bilateral involvement in 86.7% (*n* = 26), unilateral in 6.7% (*n* = 2). The most common findings were white matter injury (36.7%, *n* = 11) followed by known metabolic patterns (GA-I, Pantothenate kinase-associated neurodegeneration, or Leigh syndrome, 10.0%, *n* = 3), malformations (6.7%, *n* = 2), and term HIE (3.3%, *n* = 1); 36.7% (*n* = 11) had other structural abnormalities. Pallidal abnormalities were present in 56.7% (*n* = 17), with no significant differences between aetiology groups (Table 1).

Neurophysiological assessment demonstrated largely preserved pathways; MEPs were present and normal in all tested (*n* = 21), with normal CMCTs; SEPs were present in all tested (*n* = 24), normal in 87.5% (*n* = 21).

The sample had a median BFMDRS-Movement score of 99.00 (IQR 35.00) at baseline, with the acquired aetiology group scoring significantly higher (Table 1; Supplementary Fig. 1). The acquired group also scored significantly higher on the BFMDRS-Disability, CGI-S, GMFCS and MACS levels; COPM performance and satisfaction scores were comparable across aetiologies (Supplementary Table 1, Supplementary Fig. 1).

#### Surgery and medications

All participants underwent de novo DBS implantation except two who had revisions or replacements; all received SenSight electrodes except one (Legacy). Lead placement was satisfactory in all cases, with minor impedance abnormalities in three requiring no intervention.

At baseline, participants were receiving a median of three medications (IQR: 3.75, range: 0–6), with a significantly higher medication burden in those with acquired dystonia (Table 1). Medication classes included mainly GABAergic, glutamatergic, adrenergic and cholinergic agents.

#### LFP data characteristics

Spectral peaks were most frequently identified in the low-gamma band, with at least one peak identified in 86.4% of the analysed 20 s segments, followed by beta (77.7%) and delta (71.9%); theta and alpha peaks were less consistently detected (43.7% and 21% respectively; Fig. 2; Supplementary Table 2). Further details on the variability in peak power between channels and the longitudinal consistency of the best channel selected are in Supplementary Table 3.

**Figure 2.**
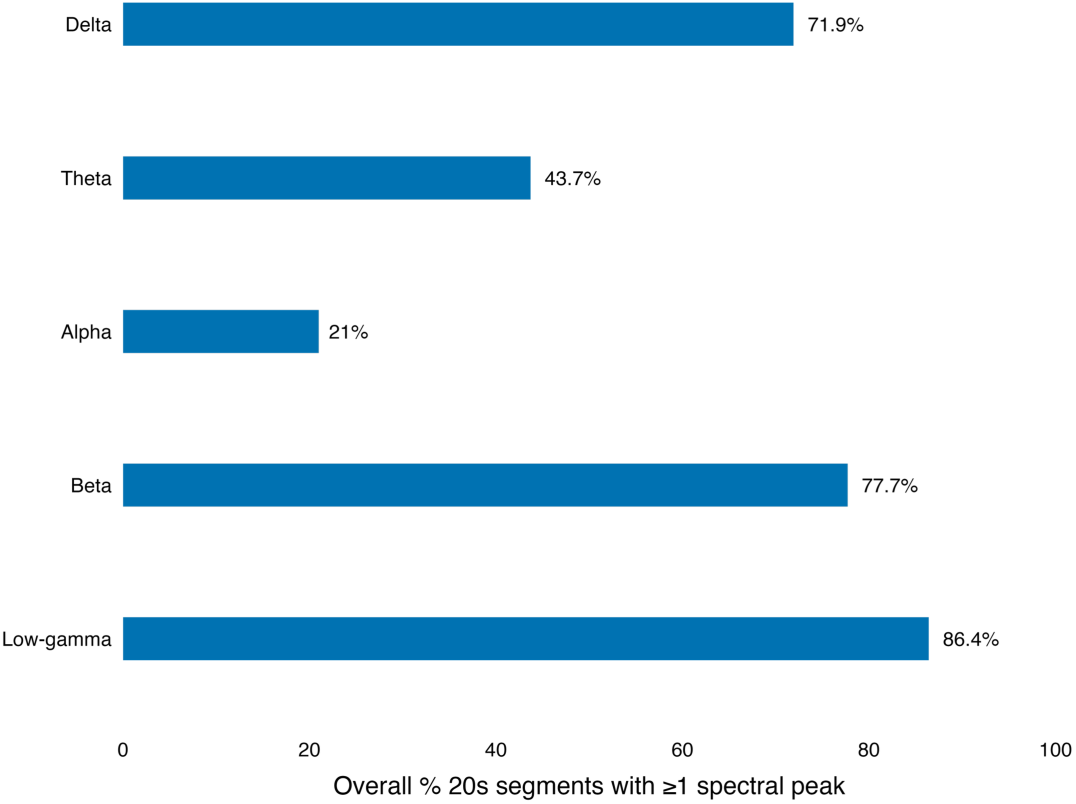
Percentage of 20 s segments with at least one peak detected. For each frequency band, reported are mean percentages across 44 recordings from 30 patients.

### Dystonia severity

#### Peak power and aperiodic exponent

Theta peak power did not significantly predict dystonia severity, as measured by BFMDRS-Movement scores (*P* = 0.47; Table 2 and Supplementary Table 6). Analyses of peak power extracted from the alpha and beta bands also revealed no association with dystonia severity (Supplementary Table 6). However, both delta and low-gamma peak power significantly predicted BFMDRS-Movement scores; for delta, lower power was associated with higher severity (FDR-corrected *P* = 0.045; Supplementary Table 6; Fig. 3). Low-gamma peak power showed a non-linear association with severity, with intermediate values associated with lower dystonia severity (FDR-corrected *P* = 0.040; Supplementary Table 6; Fig. 3).

**Table 2.** Summary results of linear mixed-effect model analyses on dystonia severity. Predictors: Theta Peak Power (best LFP channel) or Aperiodic Exponent (average across all LFP channels).

| Fixed effects | $\beta$ | 95% CI for $\beta$ (lower) | 95% CI for $\beta$ (upper) | SE | df | t | P value (uncorr.) | P value (FDR corrected) |
| --- | --- | --- | --- | --- | --- | --- | --- | --- |
| **Unadjusted** |  |  |  |  |  |  |  |  |
| Theta peak power | -6.10 | -14.02 | 1.83 | 3.90 | 34.80 | -1.56 | 0.140 | 0.233 |
| Aperiodic exponent | 7.45 | -5.94 | 21.06 | 6.73 | 33.61 | 1.11 | 0.274 | n/a |
| **Adjusted<sup>a</sup>** |  |  |  |  |  |  |  |  |
| Theta peak power | -2.13 | -7.85 | 3.78 | 2.86 | 35.46 | -0.74 | 0.473 | 0.473 |
| Aperiodic exponent | -1.25 | -11.28 | 9.17 | 5.00 | 36.80 | -0.25 | 0.790 | n/a |
| <sup>a</sup>Adjusted for pallidal involvement and medication number modelled as a spline (3 df).<br>CI = confidence intervals; SE = standard error; df = degrees of freedom; uncorr. = uncorrected; FDR = false discovery rate |  |  |  |  |  |  |  |  |

**Figure 3.**
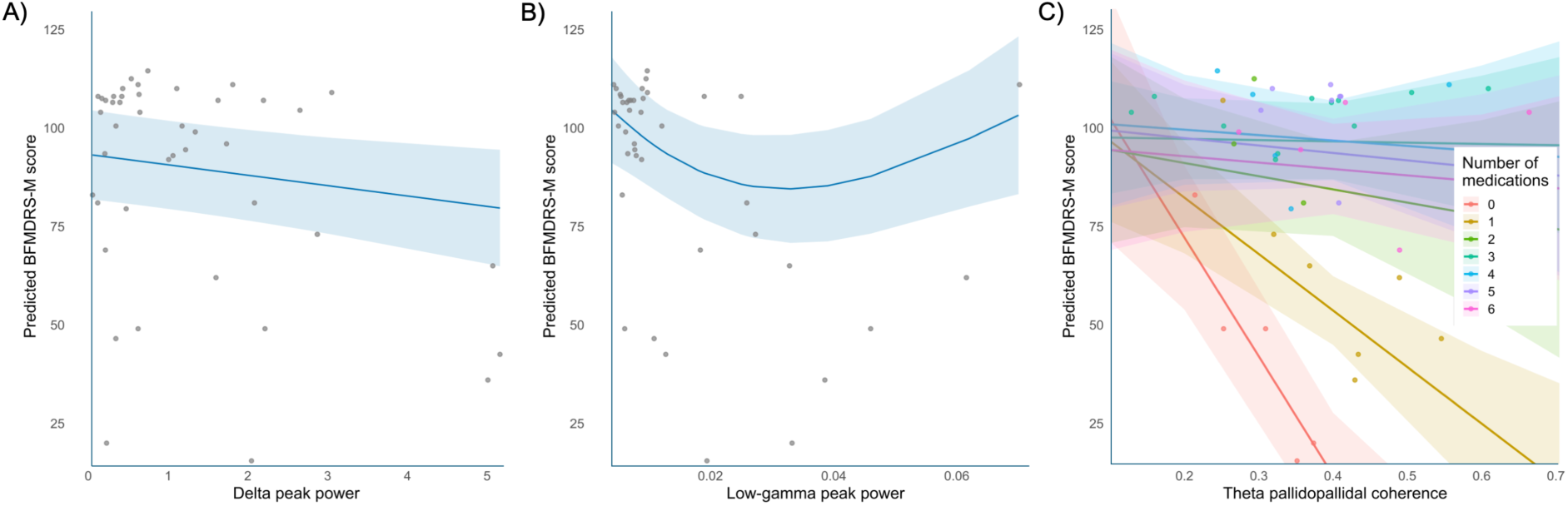
Model-predicted association between BFMDRS-M scores and (A) delta peak power, (B) low-gamma peak power, and (C) theta pallidopallidal coherence. Solid lines represent model-predicted BFMDRS-M scores from the adjusted mixed-effects models, with shaded areas indicating 95% confidence intervals. Points represent raw, unadjusted individual observations contributing to the model estimates. For C), colours indicate the number of medications.

Details on model specifications, diagnostics, and sensitivity analyses are in Supplementary Section 3.1.2. Both these and unadjusted analyses (Table 2, Supplementary Table 4) supported the conclusions of the primary analyses.

Across participants and recordings, the median knee frequency of the aperiodic exponent was 1.13 Hz (IQR = 1.40). The aperiodic exponent was not a significant predictor of dystonia severity (*P* = 0.79; Table 2; Supplementary Table 10). Unadjusted (Table 2; Supplementary Table 8) and sensitivity analyses (Supplementary Section 3.2.2) did not change these conclusions.

Consistently across all adjusted models, both covariates were significantly associated with the outcome, with pallidal involvement on MRI and higher number of medications associated with greater dystonia severity (Supplementary Tables 6 and 10; Supplementary Fig. 2).

#### Pallidopallidal coherence

Theta pallidopallidal coherence did not significantly predict BFMDRS-Movement scores (FDR-corrected *P* = 0.20; Table 3; Supplementary Tables 12 and 14). However, residual diagnostics demonstrated deviations from the expected distribution and suggested the presence of an interaction with medication number. Including an interaction term improved residual diagnostics and model fit, with a significant coherence × medication number interaction (χ²(3) = 17.02, *P* < 0.001; Table 3; Supplementary Table 16; Fig. 3).

**Table 3.** Summary results of linear mixed-effect model analyses on dystonia severity. Predictor: Theta Pallidopallidal Coherence (best LFP channel).

| Fixed effects | $\beta$ | 95% CI for $\beta$ (lower) | 95% CI for $\beta$ (upper) | SE | df | t | P value (uncorr.) | P value (FDR corrected) |
| --- | --- | --- | --- | --- | --- | --- | --- | --- |
| **Unadjusted** |  |  |  |  |  |  |  |  |
| Theta Pallidopallidal Coherence | 13.76 | -27.82 | 56.12 | 21.28 | 16.80 | 0.65 | 0.542 | 0.670 |
| **Adjusted<sup>a</sup>** |  |  |  |  |  |  |  |  |
| Theta Pallidopallidal Coherence | -31.20 | -70.05 | 7.42 | 19.51 | 31.30 | -1.60 | 0.120 | 0.200 |
| **Adjusted, with interaction term<sup>b</sup>** |  |  |  |  |  |  |  |  |
| Theta Pallidopallidal Coherence × Medication number | n/a | n/a | n/a | n/a | 3 | $\chi^2 = 17.02$ | <0.001 | <0.001 |
| <sup>a</sup>Adjusted for pallidal involvement and medication number modelled as a spline (3 df).<br><sup>b</sup>Adjusted for pallidal involvement and medication number modelled as a spline (3 df), and including the interaction term pallidopallidal coherence × medication number (3 df). The Likelihood ratio test compares the adjusted model including the coherence × medication number interaction (modelled using a 3 df spline) with the corresponding model without the interaction term.<br>CI = confidence intervals; SE = standard error; df = degrees of freedom; uncorr. = uncorrected; FDR = false discovery rate | | | | | | | | |

Pallidopallidal coherence extracted from all other frequency bands revealed similar findings (Supplementary Table 16; Supplementary Fig. 3).

### Dystonia aetiology

#### Peak power and aperiodic exponent

There was no significant association between theta peak power and the odds of having acquired rather than genetic aetiology (FDR-corrected *p* = 0.65; Table 4). The unadjusted analyses revealed similar findings (Table 4). No association with dystonia aetiology was found for peak power extracted from the delta, alpha, and beta bands (Supplementary Table 28).

**Table 4.** Results of Firth’s bias-reduced logistic regression analyses on dystonia aetiology. Predictors: Theta Peak Power (best LFP channel), Aperiodic Exponent (average across all LFP channels), or Theta Pallidopallidal Coherence (best LFP channel).

| Fixed effects | $\beta$ | SE | OR | 95% CI for OR (lower) | 95% CI for OR (upper) | P value (uncorr.) | P value (FDR corrected) |
| --- | --- | --- | --- | --- | --- | --- | --- |
| **Unadjusted** |  |  |  |  |  |  |  |
| Theta peak power | -0.60 | 0.40 | 0.54 | 0.20 | 1.14 | 0.111 | 0.185 |
| Aperiodic exponent | 5.27 | 2.29 | 194.80 | 3.21 | 63972.79 | 0.006 | n/a |
| Theta Pallidopallidal Coherence | 2.21 | 3.90 | 9.11 | 0.00 | 41037.50 | 0.569 | 0.700 |
| **Adjusted<sup>a</sup>** |  |  |  |  |  |  |  |
| Theta peak power | -0.61 | 0.56 | 0.54 | 0.11 | 1.59 | 0.326 | 0.652 |
| Aperiodic exponent<sup>b</sup> | 4.13 | 2.18 | 62.27 | 0.93 | 13560.27 | 0.056 | n/a |
| Theta Pallidopallidal Coherence | -2.91 | 4.88 | 0.05 | 0.00 | 1558.53 | 0.572 | 0.715 |
| <sup>a</sup>Adjusted for pallidal involvement and medication number, unless otherwise specified.<br><sup>b</sup>Adjusted for medication exposure only, modelled as a binary variable (0 = no medications, 1 = $\geq 1$ medication), due to estimation instability when medication number was included as a continuous predictor.<br>SE = standard error; OR = Odds Ratios; CI = confidence intervals; df = degrees of freedom; uncorr. = uncorrected; FDR = false discovery rate | | | | | | | |

In contrast, the logistic regression for low-gamma peak power failed to converge because of quasi-complete separation, precluding reliable estimation of regression coefficients. To quantify the significance of this separation, low-gamma peak power was binarised (threshold of 0.01), and Fisher’s exact test^57^ demonstrated a significant association with aetiology (P <0.001; Supplementary Table 28; Fig. 4).

**Figure 4.**
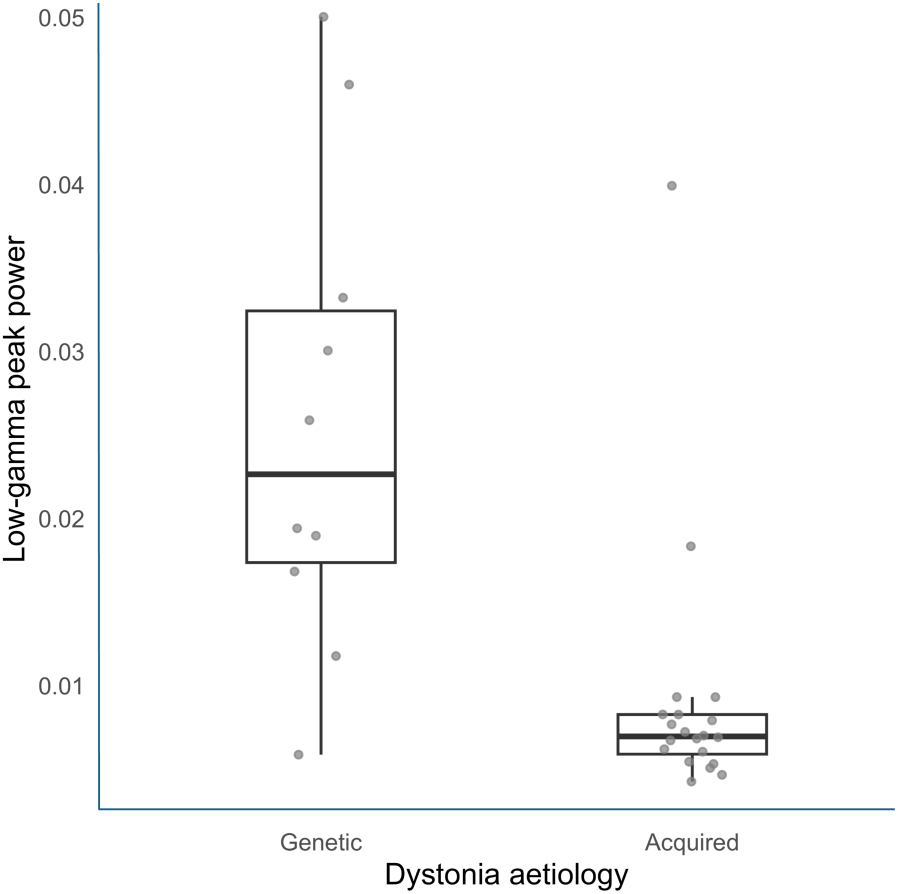
Observed association between dystonia aetiology and low-gamma peak power (best LFP channel). Grey points represent raw individual observations; boxplots depict the median and interquartile range. Low low-gamma peak power values (<0.01 threshold) occur in 18 of 20 individuals with acquired dystonia (90%), whereas high values (≥0.01) occur almost exclusively in genetic dystonia (9 of 10 individuals, 90%).

Dystonia aetiology was significantly associated with the aperiodic exponent when this was modelled independently (*P* = 0.006; Table 4). However, this association was no longer significant after adjustment for medication exposure as a binary variable (*P* = 0.056; Table 4). More granular models including medication number as a covariate were unstable due to quasi-complete separation.

#### Pallidopallidal coherence

There was no significant association between pallidopallidal coherence in any frequency band and the odds of having acquired rather than genetic aetiology (theta: FDR-corrected *P* = 0.71; Table 4; other frequency bands: Supplementary Table 29). Conclusions remained unchanged in unadjusted analyses (Table 4; Supplementary Table 29).

Consistently across all adjusted models, medication number was significantly associated with dystonia aetiology but pallidal involvement on MRI was not (Supplementary Fig. 4).

### Control analyses

Control analyses using power extracted from a 3-Hz narrow band centered on the spectral peak continued to demonstrate significant associations between delta and low-gamma power and dystonia severity; however, neither remained significant after FDR correction (Supplementary Table 18).

Analyses using average peak power across all LFP channels continued to support significant associations between severity and both delta peak power and pallidopallidal coherence across all frequency bands, but not low-gamma peak power (Supplementary Tables 20, 22).

Low-gamma power remained significantly associated with dystonia aetiology when this was extracted from a 3-Hz band or averaged across all channels (Supplementary Tables 30-32).

Delta peak power, low-gamma peak power and coherence in all bands remained significant predictors of severity after adjustment for dystonia aetiology (Supplementary Table 24).

Control models including dystonia severity as an additional covariate when predicting aetiology from low-gamma peak power were not identifiable due to quasi-complete separation, precluding estimation of the independent association with aetiology beyond the potentially confounding effect of severity.

Sensitivity analyses excluding four recordings (two acquired during sleep, two from a participant with detected epileptiform discharges) yielded results consistent with the primary analyses (Supplementary Table 26).

## Discussion

Bidirectional DBS systems offer the opportunity to identify neurophysiological biomarkers and work towards developing adaptive, closed-loop DBS paradigms for neurological and neuropsychiatric disorders. In Parkinson’s disease, sub-thalamic beta oscillations are already being used to guide adaptive DBS.^58^ In dystonia, the identification of equivalent physiomarkers remains an ongoing challenge, complicated by the fact that the clinical effects of DBS programming often take days to weeks to manifest. Whilst adult studies have highlighted the importance of pallidal low-frequency oscillations in dystonia, their specificity, reliability, and generalisability across age groups and dystonia aetiologies remain poorly characterised. These represent important considerations in the development of robust, objective LFP-guided stimulation protocols.

To our knowledge, this study reports the largest dataset to date of pallidal LFP recordings in children and young people with refractory dystonia of both genetic and acquired origin. Results indicate that pallidal LFP markers (delta peak power, low-gamma peak power and pallidopallidal coherence) are associated with dystonia severity in children. These show different characteristics from those reported in adults.

### Delta and low-gamma peak power are significant predictors of dystonia severity

We observed frequent detectable spectral peaks in the delta, beta, and low-gamma frequency bands. Delta and low-gamma peak power were significant predictors of dystonia severity, as measured by the BFMDRS-M. This association was independent of medication burden, structural GPi involvement on MRI, and dystonia aetiology. Conversely, theta and alpha peaks were only present during a minority of the recording time, and their power was not associated with dystonia severity.

Our results broadly align with previous literature in supporting the relevance of low-frequency oscillations in dystonia, but we specifically identify the 1–4 Hz (delta) band, rather than the more commonly reported 4–10 Hz in adults.^10^ Previous reports of pallidal LFPs in paediatric patients, limited to a few case-studies and small series, have shown inconsistent findings regarding the most prominent peak frequencies and associations with symptom severity: some implicate the delta band;^20,27^ others report alpha^19,29^ or beta.^28,29^ In this study, lower delta power was associated with higher severity suggesting that during development, the neurophysiological substrate of dystonia severity may be characterised by a distinct pallidal spectral profile.

Although animal studies indicate that basal ganglia activity undergoes substantial changes in spontaneous activity during development, the longitudinal profile of healthy subcortical oscillatory physiology in humans remains largely unknown.^59–62^ In the cortex, low-frequency activity is prominent during childhood and becomes progressively less dominant with age.^63,64^ Given that the cortex is a major driver of activity within basal ganglia nuclei, subcortical oscillatory activity may follow a similar trajectory. The observed association between greater delta power and lower dystonia severity may indicate that preserved developmentally typical low-frequency dynamics support the organisation of motor networks during maturation, contributing to a healthier functional state. Increased delta activity is also observed during GABAergic anaesthesia,^65,66^ raising the possibility that greater delta power may reflect a relatively more inhibited network state associated with less severe dystonia. Longitudinal LFP studies are needed to understand the developmental trajectories of pathological oscillatory activity in the basal ganglia.

We also observed a significant association between low-gamma (30–45 Hz) peak power and dystonia severity, with an optimal, intermediate range of activity associated with lower severity and both low and high gamma levels predictive of greater severity. Gamma activity, which is related to the time constants of GABA_A_-mediated signalling^67^ – abundant in the GPi – is the most prominent resting basal ganglia rhythm in healthy mammals^68,69^ and emerges following restoration of dopaminergic drive in people with Parkinson’s disease,^70^ supporting its role as a physiologically healthy feature of resting basal ganglia circuits. Additionally, transient basal ganglia gamma changes (>30 Hz) occur during movement execution and inhibition and sustained, finely tuned oscillations (60–90 Hz) have been observed in prokinetic clinical states.^54–56^ In children with active dystonia, a high prevalence of gamma peaks might reflect preserved physiological recruitment of pallidal circuits, with intermediate low-gamma activity potentially reflecting an optimal level of network engagement supporting appropriately timed pallidal inhibitory output to downstream structures. This would be consistent with the idea that an optimal temporal pattern of activity creates the conditions for balanced and flexible excitatory and inhibitory dynamics supporting healthy movement^72,73^ and with computational studies postulating that gamma can emerge when the basal ganglia operate within an appropriate dynamic regimen, with reduced or excessive gamma reflecting pathological network states.^74^

The association between low-gamma and dystonia severity also raises the possibility that gamma is not only subserving communication but also plasticity-driven motor consolidation processes during development. Gamma activity has been shown to align neuronal firing within the critical temporal windows relevant for spike-timing-dependent plasticity,^20,67,75,76^ a process shaped by GABAergic signalling.^67,77^ This is relevant to dystonia, where maladaptive plasticity has been proposed to contribute to consolidation of abnormal motor representations.^73,78^ Given the wider role of the basal ganglia in reinforcement-based learning and action-sequence acquisition,^79^ and the predominantly GABAergic nature of GPi output, an optimal range of low-gamma activity could potentially support the development and maintenance of healthier motor representations during development, whereas either reduced or excessive low-gamma activity might disrupt this balance. However, direct evidence linking 30–45 Hz activity to Hebbian plasticity within the GPi is currently lacking, so this interpretation remains speculative.

Recordings were acquired during clinical appointments, typically during rest, or during spontaneous activities such as using a tablet, eating, etc. Therefore, we cannot completely exclude the possibility that the findings represent transient physiological activity associated with voluntary or involuntary movement^54–56^ or alertness levels.^80–82^

However, previous studies have observed voluntary movement-related gamma event-related synchronisation in the contralateral GPi of dystonia patients in the 60–80 Hz range,^83,84^ and independently of dystonic symptoms,^84^ which is higher than the 30–45 Hz range here. Changes in alertness are also unlikely to solely explain our findings, as sensitivity analyses excluding sleeping patients did not alter results.

Collectively, these findings suggest that peak power signatures identified in adult cohorts (i.e. 4-10 Hz oscillations) may not fully generalise to the developing brain, where the disruption of delta and low-gamma operating regimes appears to be more closely related to symptom severity.

### Pallidopallidal coherence predicts dystonia severity in interaction with medication load

Across frequency bands, pallidopallidal coherence also significantly predicted dystonia severity in interaction with medication load. In unmedicated patients and those receiving monotherapy, lower coherence was associated with greater severity; this association no longer applied in people on polytherapy (two or more medications). Although these findings should be considered exploratory due to the number of model terms included, an interaction was strongly supported and the effect was consistent across frequency bands, suggesting a potentially meaningful finding.

Functional homotopy, the spontaneous synchronisation of homologous regions across hemispheres, is a fundamental characteristic of brain architecture, with robust connectivity in the basal ganglia, somatosensory, motor and premotor cortices.^85^ This reflects both structural and functional connectivity and is thought to support integrated network activity.^85–90^ Impaired homotopic connectivity has been described in several disorders.^85,90,91^ Evidence for pallidopallidal coherence in dystonia is scarce,^11^ with a single study examining its relationship with symptom severity and reporting an opposite association^12^; in contrast, reduced homotopic connectivity at the cortical level has been reported in focal dystonias.^11,92–95^

In our cohort, lower pallidopallidal coherence consistently reflected greater severity. Although the limited literature precludes a clear mechanistic interpretation, this may indicate adoption of alternative, maladaptive signalling pathways within the motor network. The variability in severity by medication load despite similarly high coherence levels may be due to differences in the underlying disease state prompting treatment; alternatively, medication load may act as a stratification factor for disorder complexity or symptom control. The monotherapy association was not attributable to a single medication or medication class.

### Low-gamma peak power is a significant predictor of aetiology, but the potential confounding effect of severity cannot be excluded

Previous studies have suggested that pallidal neuronal firing patterns^96^ and brain glucose metabolism^97^ may differ according to dystonia aetiology. However, the extent to which LFP features vary across aetiologies is relatively unexplored, with previous studies limited by small sample sizes and inclusion of few individuals with acquired dystonias.^10,11^ In this study, direct statistical comparison demonstrated broadly comparable LFP characteristics across genetic and acquired dystonias, with no group differences in delta, theta, alpha, and beta peak power, aperiodic exponent and pallidopallidal coherence. We nevertheless observed significantly lower low-gamma peak power in the acquired group. Unfortunately, statistical considerations prevented us from determining whether this association was independent of dystonia severity. This is relevant given that severity was significantly higher in the acquired group, which likely reflects selection bias inherent to our tertiary service, where DBS is considered for genetic dystonias across a broad range of severities but is typically indicated for more severe, treatment-refractory acquired cases. Replication in larger, independently recruited cohorts with different sampling strategies is required to establish whether low-gamma power is indeed associated with dystonia aetiology. Taken together, our findings reinforce previous observations that pallidal physiomarkers may primarily reflect disease expression rather than the underlying mechanisms driving dystonia pathogenesis,^5,10,11^ with the isolated low-gamma difference requiring cautious interpretation given potential confounding by dystonia severity.

### Additional findings and methodological considerations

Our primary analyses examined peak power extracted from each individual’s “best channel” (i.e., the LFP pair displaying the highest and most frequent peaks). A series of control analyses confirmed that the observed associations persisted, but in an attenuated form, when power was averaged over a 3-Hz narrow band around the peak, closely reproducing results from a previous study^12^ and supporting the use of power at the peak centre as the most sensitive measure. For delta, the association with severity also remained significant when considering the average peak power across all 12 LFP pairs, whereas the low-gamma association did not. This suggests that the low-gamma physiomarker may reflect a more locally restricted process, consistent with the functional segregation of basal ganglia territories and the spatial specificity of pallido-subthalamic connections associated with optimal DBS outcomes.^98^ For delta, the persistence of the association across LFP pairs may reflect the spatially broader neuronal assemblies engaged in delta oscillations^99^ or greater consistency of delta activity across territories. Multi-site (e.g., EEG and LFP) recordings would be needed to determine whether different oscillation frequencies reflect communication at different spatial scales within the dystonia network.^99,100^

Although beta peaks were frequently present, these were not associated with severity or aetiology, warranting further investigation of their clinical correlates, including the potential effect of medications.^10,101^

In unadjusted models, the aperiodic exponent was significantly associated with dystonia aetiology, but this association was no longer significant after adjusting for medication load. This effect is biologically plausible, given evidence that the aperiodic exponent tends to be modulated by medications in general^102^ and interventions affecting basal ganglia activity in movement disorders in particular.^103–105^ These findings highlight the importance of parameterising the power spectrum to disentangle oscillatory and non-oscillatory activity and controlling for medication effects.

Regarding confounders, the presence of structural GPi abnormalities on MRI was associated with greater dystonia severity, providing evidence that the structural integrity of pallidal tissue contributes to dystonia’s clinical expression. Structural GPi involvement occurred in similar proportion across aetiological groups in our cohort.

### Limitations and future directions

Despite representing one of the largest reported cohorts of LFP data in dystonia, limitations include the relatively small sample size, inherent to invasive recordings; this limits statistical power and the number of covariates that can be reliably studied. Both our genetic and acquired groups were heterogeneous; although representative of the clinical population, this precluded direct comparison between specific genetic aetiologies. This should be investigated in larger studies. One-year follow-up data was only available for 14 patients; however, this was not considered a limitation as the follow-up data increased power for the severity analyses by allowing changes in this state-level marker, while averaging across timepoints in the aetiology analysis mitigated state-related variability to facilitate identification of this stable, trait-level marker. In our pallidopallidal coherence analyses, the presence of non-linear relationships and interactions inevitably increased model parameters over recommended numbers; results should be treated as exploratory, with replication in larger cohorts recommended. Recordings were obtained during clinical appointments without systematic assessment of voluntary or involuntary movement or arousal levels beyond awake/asleep state; future studies should incorporate objective monitoring. Comparative studies including other hyperkinetic disorders will be essential to establish the specificity of these physiomarkers to dystonia. The cross-sectional nature of the study precludes determining whether the observed LFP patterns represent a cause or an epiphenomenon of dystonia. Longitudinal studies defining developmental trajectories of cortico-basal ganglia LFP activity may provide further insights. This study focused on associations with the BFMDRS as a severity measure. It remains to be established whether any long-term treatment-related changes in BFMDRS correlate with changes in LFP markers. For the development of future adaptive DBS systems, it will be important to determine whether LFP changes relate to more rapid fluctuations in severity and which behavioural or physiological measures are most sensitive to these changes.

## Conclusion

Pallidal LFP markers are associated with dystonia severity in children but show different characteristics from previously-reported adult cohorts. These findings support the potential of pallidal LFPs to characterise dystonia across the lifespan and highlight the need for longitudinal studies of their developmental and clinical trajectories. Defining these trajectories will be essential for designing robust LFP-guided stimulation protocols.

## Supporting information

Supplementary Material - Brainsense_ecg_removal method description and synthetic validation

## Data availability

De-identified data supporting the findings of this study are available upon reasonable request.

## Acknowledgements

We would like to acknowledge Dr John Hodsoll for his statistical advice, Dr Michael Everett Rule for reviewing the brainsense_ecg_removal function code, and Elena Cioffi and Anna Hutber for their assistance with the initial study set-up.

## Funding

This project is supported by a grant from King’s Health Partners Centre for Translational Medicine Pilot Fund. Dr. McClelland is also supported by a Clinician Scientist Fellowship from the Medical Research Council UK (MRP0068681) and by a Continuation Funding grant from the Rosetrees Trust (CF-2021-2-112).

## Competing interests

The authors report no competing interests.

## Supplementary material

Supplementary material is available at *Brain* online.

## Notes

### Competing Interest Statement

The authors have declared no competing interest.

### Author Declarations

Research Ethics Committee of East Midlands - Nottingham 1 gave ethical approval for this work (REC: 23/EM/0035, IRAS ID: 317454, latest amendment: 03/02/2025).

### Summary of Updates

Spelling error in one of the author's names has been corrected. No other changes.

