## Supplementary Material for "Pallidal Local Field Potentials markers of dystonia aetiology and severity in children"

*4.5 Control analyses. Predictor: Pallidopallidal coherence (average across all LFP channels) 21*

### Supplementary Section 1. Methods

#### Clinical Global Impression Scale

Reproduced from Guy, W. (1976). *ECDEU assessment manual for psychopharmacology*. US Department of Health, Education, and Welfare, Public Health Service, Alcohol, Drug Abuse, and Mental Health Administration, National Institute of Mental Health, Psychopharmacology Research Branch, Division of Extramural Research Programs.

##### Clinical Global Impression (CGI) Scale

**1. SEVERITY OF DYSTONIA**

Considering your total clinical experience with this particular population, how dystonic is the patient at this time?

0 = Not assessed
1 = Normal, not dystonic at all
2 = Borderline dystonic
3 = Mildly dystonic
4 = Moderately dystonic
5 = Markedly dystonic
6 = Severely dystonic
7 = Among the most extremely dystonic patients

*The next items may be omitted at the initial assessment by marking "not assessed"*

**2. GLOBAL IMPROVEMENT**

Rate total improvement whether or not, in your judgement, it is due entirely to DBS treatment.
Compared to his/her dystonia at baseline (pre-DBS), how much has he/she changed?

| | |
|---|---|
| 0 = Not assessed<br>1 = Very much improved<br>2 = Much improved<br>3 = Minimally improved | 4 = No change<br>5 = Minimally worse<br>6 = Much worse<br>7 = Very much worse |

3. **EFFICACY INDEX** – Rate this item on the basis of DBS EFFECT ONLY

| | |
|---|---|
| 0 = Not assessed<br>1 = Very much improved<br>2 = Much improved<br>3 = Minimally improved | 4 = No change<br>5 = Minimally worse<br>6 = Much worse<br>7 = Very much worse |

**4. THERAPEUTIC VS SIDE EFFECT MATRIX**

| | | SIDE EFFECTS | | | |
|---|---|---|---|---|---|
| | | None | Do not significantly interfere with patient's functioning | Significantly interferes with patient's functioning | Outweighs therapeutic effects |
| THERAPEUTIC EFFECT | MARKED – Vast improvement. Complete or nearly complete remission of all symptoms. | 01 | 02 | 03 | 04 |
| | MODERATE – Decided improvement. Partial remission of symptoms. | 05 | 06 | 07 | 08 |
| | MINIMAL – Slight improvement which doesn't alter status of care of patient | 09 | 10 | 11 | 12 |
| | UNCHANGED OR WORSE | 13 | 14 | 15 | 16 |
| | Not Assessed = 00 | | | | |

### Supplementary Section 2. Results: Population description

#### Non-motor neurological comorbidities

As documented in clinical notes, non-motor neurological comorbidities for the study sample included Developmental Delay / Learning Difficulties (n = 8), history of seizures (confirmed or suspected, n = 5), vision or hearing abnormalities (n = 9), Autism Spectrum Disorder (n = 3), Attention Deficit Hyperactivity Disorder (n = 4), anxiety disorders (n = 2), other neuromuscular abnormalities (n = 2).

#### Supplementary Table 1. Baseline scores for clinical scales administered as part of presurgical assessment for the whole sample and for each aetiology group, with statistical comparison.

| | **Whole sample (*n* = 30)** | **Aetiology: Genetic (*n* = 10)** | **Aetiology: Acquired (*n* = 20)** | **Statistic** | **p-value** |
|---|---|---|---|---|---|
| CGI, median (IQR) | 5.50 (1.00) | 4.25 (1.88) | 6.00 (1.12) | 52.0 | 0.033* |
| BFMDRS-Disability score, median (IQR) | 28.00 (13.00)<br>*n* = 29 | 16.00 (8.00)<br>*n* = 9 | 29.00 (3.00) | 30.0 | 0.004** |
| GMFCS level, median (IQR) | 5.00 (1.00)<br>*n* = 29 | 2.00 (2.50)<br>*n* = 9 | 5.00 (0.25) | 28.5 | 0.001** |
| MACS level, median (IQR) | 5.00 (1.25)<br>*n* = 28 | 2.00 (2.00)<br>*n* = 9 | 5.00 (0.00)<br>*n* = 19 | 16.5 | <0.001*** |
| COPM-Performance, median (IQR) | 2.90 (1.95)<br>*n* = 27 | 1.80 (1.12)<br>*n* = 8 | 3.20 (1.90)<br>*n* = 19 | 47.0 | 0.129 |
| COPM-Satisfaction, median (IQR) | 2.20 (1.65)<br>*n* = 27 | 1.15 (1.25)<br>*n* = 8 | 2.20 (1.60)<br>*n* = 19 | 41.5 | 0.070 |
| CPCHILD total, mean (SD) | 38.94 (10.16)<br>*n* = 21 | 42.90 (15.41)<br>*n* = 2 | 38.53 (9.97)<br>*n* = 19 | $t(1.08) = 0.39$ | 0.757 |

CGI, Clinical Global Impression; BFMDRS, Burke-Fahn-Marsden Dystonia Rating Scale; GMFCS, Gross Motor Function Classification System; MACS, Manual Ability Classification System; COPM, Canadian Occupational Performance Measure; CPCHILD, Caregiver Priorities and Child Health Index of Life with Disabilities. The supporting sample size for each measure is indicated in the column headers unless otherwise specified.

#### Supplementary Figure 1. Distribution of clinical scale scores administered as part of presurgical assessment, for the whole sample and for each aetiology group.

BFMDRS, Burke-Fahn-Marsden Dystonia Rating Scale; CGI, Clinical Global Impression – Severity; GMFCS, Gross Motor Function Classification System; MACS, Manual Ability Classification System; COPM, Canadian Occupational Performance Measure; CPCHILD, Caregiver Priorities and Child Health Index of Life with Disabilities. The supporting sample size for each measure is indicated in the column headers unless otherwise specified.

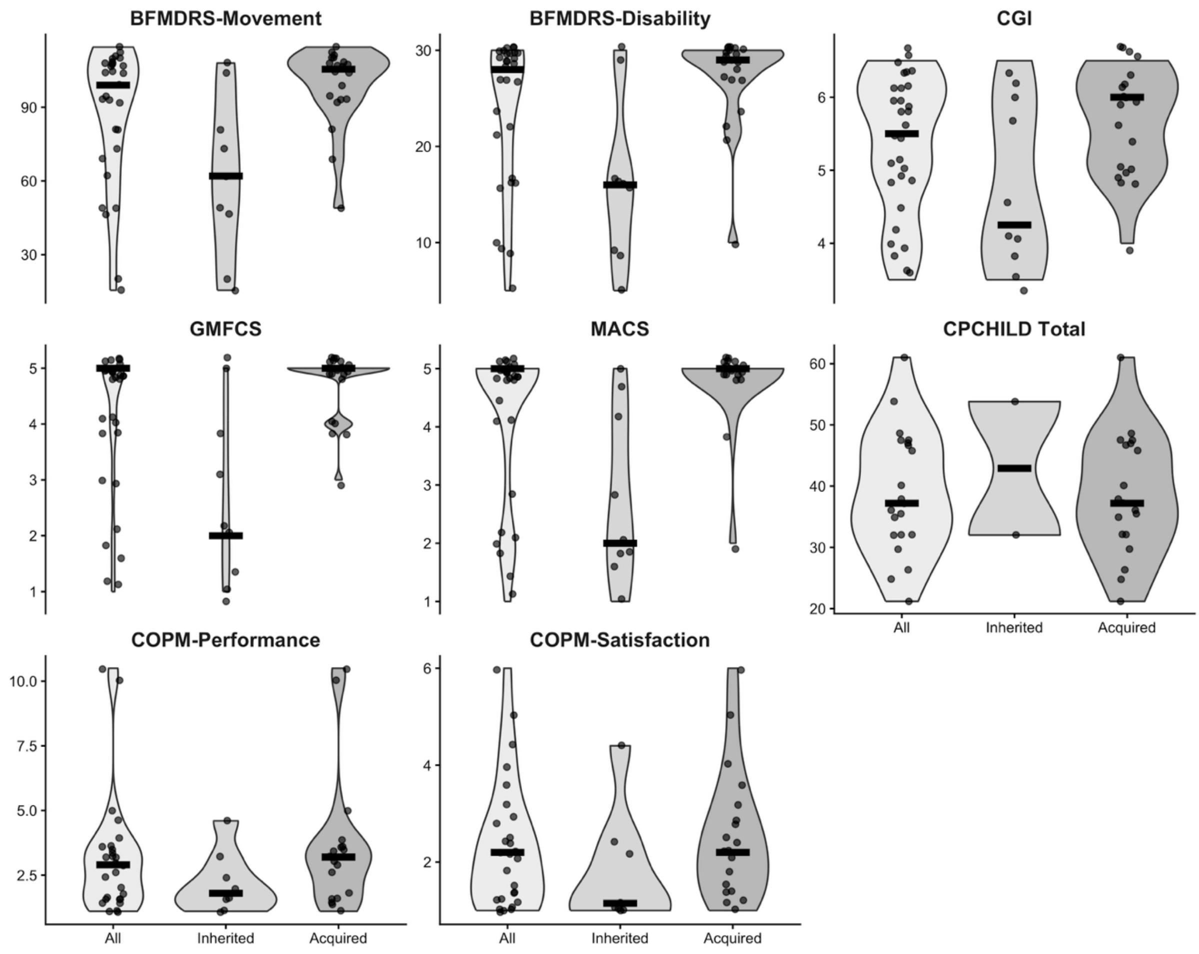

Supplementary Table 2. Percentage of 20 s segments with 0, 1 or 2 or more peaks detected. Reported are mean (SD) percentages across 44 recordings from 30 patients.

| | % segments with 0 peaks | % segments with 1 peak | % segments with >=2 peaks |
|---|---|---|---|
| **Delta** | 28.1 (18.9) | 61.8 (14.3) | 10.1 (10.7) |
| **Theta** | 56.3 (29.0) | 40.7 (26.2) | 3.0 (3.4) |
| **Alpha** | 79.1 (20.0) | 19.8 (18.4) | 1.2 (2.9) |
| **Beta** | 22.4 (23.3) | 35.9 (12.6) | 41.7 (26.6) |
| **Gamma** | 13.6 (18.6) | 30.2 (12.5) | 56.2 (23.4) |

Supplementary Table 3. Variability in peak power and longitudinal consistency of best channel index.

Coefficient of variation represents the variability in weighted average peak power between LFP channels within each frequency band. For participants with 12-month follow-up ($n$ = 14), the table also reports the number ($n$) and percentage (%) of participants in whom the best channel index remained unchanged between baseline and 12 months.

| | Coefficient of Variation, mean (SD) across recordings | Repeated-measure consistency of best channel selection (n) | Repeated-measure consistency of best channel selection (%) |
|---|---|---|---|
| **Delta** | 0.43 (0.19) | 6 | 42.86 |
| **Theta** | 0.49 (0.19) | 2 | 14.28 |
| **Alpha** | 0.61 (0.26) | 2 | 14.28 |
| **Beta** | 0.36 (0.15) | 5 | 35.71 |
| **Gamma** | 0.22 (0.16) | 3 | 21.42 |

### Supplementary Section 3. Results for dystonia severity: Mixed-effect models

#### 3.1 Predictor: Peak power (extracted from best LFP channel)

##### 3.1.1 Unadjusted models

**Supplementary Table 4.** In-detail fixed effect results for linear mixed-effect model analyses on dystonia severity. Predictor: peak power (best LFP channel). Covariates: none (unadjusted analyses).

| Fixed effects | β | 95% CI for β (lower) | 95% CI for β (upper) | *SE* | df | t | *P* value *(uncorr.)* | *P* value *(FDR corrected)* |
|---|---|---|---|---|---|---|---|---|
| **Delta** | | | | | | | | |
| Intercept | 90.43 | 80.63 | 100.30 | 5.13 | 31.79 | 17.64 | <0.001 | |
| Peak power | -3.12 | -5.06 | -1.11 | 0.98 | 12.78 | -3.19 | 0.002 | *0.010* |
| **Theta** | | | | | | | | |
| Intercept | 90.55 | 79.24 | 101.49 | 5.65 | 34.02 | 16.03 | <0.001 | |
| Peak power | -6.10 | -14.02 | 1.83 | 3.90 | 34.80 | -1.56 | 0.140 | 0.233 |
| **Alpha** | | | | | | | | |
| Intercept | 87.85 | 77.05 | 98.55 | 5.47 | 33.97 | 16.06 | <0.001 | |
| Peak power | -7.37 | -28.37 | 14.15 | 10.78 | 15.26 | -0.68 | 0.498 | *0.498* |
| **Beta** | | | | | | | | |
| Intercept | 88.41 | 77.99 | 98.83 | 5.33 | 32.77 | 16.57 | <0.001 | |
| Peak power | -16.48 | -41.54 | 9.83 | 13.18 | 13.08 | -1.25 | 0.215 | *0.268* |
| **Gamma** | | | | | | | | |
| Intercept | 99.06 | 88.01 | 110.38 | 5.66 | 36.55 | 17.51 | <0.001 | |
| Peak power | n/a | n/a | n/a | n/a | 2 | $\chi^2$ = 9.31[a] | *0.009* | *0.022* |

[a] $\chi^2$ for gamma peak power compares a model without peak power to one with peak power modelled as a natural spline (2 df). The relationship between peak power and BFMDRS-M was modelled using a natural spline (2 df) as this significantly improved both model fit ($\chi^2(1)$ = 9.45, ***P*** = 0.002) and the distribution of residuals relative to a linear term. Uncorr. = uncorrected

**Supplementary Table 5. In-detail random effect results for linear mixed-effect model analyses on dystonia severity.** Predictor: peak power (best LFP channel). Covariates: none (unadjusted analyses).

| Random effects | Variance | SD | 95% CI for SD (lower) | 95% CI for SD (upper) |
|---|---|---|---|---|

| | | | | |
|---|---|---|---|---|
| **Delta** | | | | |
| Participant Intercept | 725.76 | 26.94 | 19.94 | 34.07 |
| Residual | 25.91 | 5.09 | 3.11 | 7.12 |
| **Theta** | | | | |
| Participant Intercept | 739.56 | 27.19 | 19.91 | 34.57 |
| Residual | 38.81 | 6.23 | 3.79 | 8.69 |
| **Alpha** | | | | |
| Participant Intercept | 771.76 | 27.78 | 20.37 | 35.32 |
| Residual | 40.96 | 6.40 | 3.88 | 8.97 |
| **Beta** | | | | |
| Participant Intercept | 763.21 | 27.63 | 20.43 | 35.0 |
| Residual | 38.79 | 6.23 | 3.80 | 8.7 |
| **Gamma** | | | | |
| Participant Intercept | 505.48 | 22.48 | 15.66 | 29.07 |
| Residual | 61.99 | 7.87 | 4.60 | 11.30 |

##### 3.1.2 Adjusted models

**Model specifications and diagnostics.**

Results of the fixed effects indicated that theta peak power did not significantly predict dystonia severity, as measured by BSMDRS-Movement scores ($P = 0.47$; Table 2 and Supplementary Table 6). Medication number was modelled using a natural spline (3 degrees of freedom) due to evidence of non-linearity (likelihood ratio test comparing models with linear versus spline term: $\chi^2(2) = 14.58$, $P < 0.001$). DHARMa diagnostics showed no significant deviation from the expected residual distribution. Levene's test indicated that residual variance differed between groups with or without pallidal involvement ($P < 0.001$). A sensitivity analysis using a heteroskedastic Gaussian model showed that modelling residual variance as a function of pallidal involvement did not significantly improve model fit (likelihood ratio test: $\chi^2(1) = 0.66$, $P = 0.41$), and the estimated effect of theta peak power remained non-significant ($\beta = -1.49$, SE = 2.71, $P = 0.58$).

Both delta and low-gamma peak power significantly predicted BFMDRS-Movement scores; for the delta band, lower power was associated with higher dystonia severity (FDR-corrected $P = 0.045$; Supplementary Table 6; Fig. 3). Low-gamma peak power was modelled using a natural spline (2 df) following evidence of non-linearity (likelihood ratio test comparing models with linear and spline terms: $\chi^2(1) = 9.45$, $P = 0.002$), with significant improvements in residuals' distributions. The resulting non-linear association is depicted in Fig. 3, with intermediate gamma peak power values associated with lower dystonia severity (likelihood ratio test for inclusion of the 2df spline term: $\chi^2 = 9.56$, $P = 0.008$; FDR-corrected $P = 0.040$; Supplementary Table 6).

For both delta and gamma peak power models, residuals followed the expected distribution, but Levene's test indicated heterogeneity of variance across pallidal involvement groups. Sensitivity analyses accounting for heteroskedasticity did not change the conclusions for either predictor (delta peak power $\beta = -3.85$, SE = 1.17, $P = 0.001$; likelihood ratio test for improved model fit compared with the homoskedastic model: $\chi^2(1) = 2.37$, $P = 0.12$; gamma peak power: heteroskedasticity modelling improved model fit, $\chi^2(1) = 5.80$, $P = 0.01$, without altering the results, $\chi^2(2) = 14.62$, $P < 0.001$ for gamma peak power modelled using a 2 df spline).

**Supplementary Table 6.** In-detail fixed effect results for linear mixed-effect model analyses on dystonia severity. Predictor: peak power (best LFP channel). Covariates: pallidal involvement and medication number.

| Fixed effect | β | 95% CI for β (lower) | 95% CI for β (upper) | *SE* | df | t | *P* value (uncorr.) | *P* value (*FDR corrected)* |
|---|---|---|---|---|---|---|---|---|
| **Delta** | | | | | | | | |
| Intercept | 44.75 | 32.27 | 57.70 | 6.53 | 31.77 | 6.86 | <0.001 | |
| Peak power | -2.64 | -4.83 | -0.38 | 1.09 | 14.40 | -2.41 | 0.018 | *0.045* |
| Pallidal involvement | 14.71 | 3.34 | 25.59 | 5.62 | 25.13 | 2.62 | 0.012 | |
| Medication number | n/a | n/a | n/a | n/a | 3 | $\chi^2$ = 30.56[a] | <0.001 | |
| **Theta** | | | | | | | | |
| Intercept | 42.24 | 28.34 | 56.31 | 7.07 | 30.05 | 5.97 | <0.001 | |
| Peak power | -2.13 | -7.85 | 3.78 | 2.86 | 35.46 | -0.74 | 0.473 | 0.473 |
| Pallidal involvement | 12.72 | 0.63 | 24.40 | 5.97 | 25.00 | 2.13 | 0.041 | |
| Medication number | n/a | n/a | n/a | n/a | 3 | $\chi^2$ = 33.26[a] | <0.001 | |
| **Alpha** | | | | | | | | |
| Intercept | 41.27 | 28.12 | 54.63 | 6.66 | 30.35 | 6.20 | <0.001 | |
| Peak power | -9.16 | -28.87 | 10.86 | 9.85 | 23.63 | -0.93 | 0.370 | *0.462* |
| Pallidal involvement | 14.42 | 2.48 | 25.93 | 5.89 | 26.01 | 2.45 | 0.020 | |
| Medication number | n/a | n/a | n/a | n/a | 3 | $\chi^2$ = 34.26[a] | <0.001 | |
| **Beta** | | | | | | | | |
| Intercept | 41.51 | 28.26 | 54.63 | 6.64 | 31.11 | 6.25 | <0.001 | |
| Peak power | -15.79 | -41.70 | 11.23 | 13.20 | 14.92 | -1.20 | 0.249 | *0.415* |
| Pallidal involvement | 14.19 | 2.28 | 25.64 | 5.86 | 26.16 | 2.42 | 0.022 | |
| Medication number | n/a | n/a | n/a | n/a | 3 | $\chi^2$ = 33.71 [a] | <0.001 | |
| **Gamma** | | | | | | | | |
| Intercept | 52.69 | 37.60 | 67.72 | 7.42 | 33.51 | 7.11 | <0.001 | |
| Peak power | n/a | n/a | n/a | n/a | 2 | $\chi^2$ = 9.56 | 0.008 | *0.040* |
| Pallidal involvement | 7.56 | -3.08 | 18.56 | 5.52 | 25.21 | 1.37 | 0.163 | |
| Medication number | n/a | n/a | n/a | n/a | 3 | $\chi^2$ = 34.70 [a] | <0.001 | |

[a] $\chi^2$ for medication number compares a model without the medication term to one including medication number modelled as a natural spline (with degrees of freedom equal to the number reported).
[b] $\chi^2$ for gamma peak power compares a model without peak power to one with peak power modelled as a natural spline (2 df). The relationship between peak power and BFMDRS-M was modelled using a natural spline (2 df) as this significantly improved both model fit ($\chi^2(1) = 9.45$, ***P*** = 0.002) and the distribution of residuals relative to a linear term.

**Supplementary Table 7. In-detail random effect results for linear mixed-effect model analyses on dystonia severity.** Predictor: peak power (best LFP channel). Covariates: pallidal involvement and medication number.

| Random effect | Variance | SD | 95% CI for SD (lower) | 95% CI for SD (upper) |
|---|---|---|---|---|
| **Delta** | | | | |
| Participant Intercept | 186.39 | 13.65 | 9.04 | 17.98 |
| Residual | 34.90 | 5.91 | 3.47 | 8.42 |
| **Theta** | | | | |
| Participant Intercept | 202.50 | 14.23 | 9.24 | 18.91 |
| Residual | 42.39 | 6.51 | 3.88 | 9.14 |
| **Alpha** | | | | |
| Participant Intercept | 194.85 | 13.96 | 9.08 | 18.44 |
| Residual | 43.76 | 6.62 | 3.92 | 9.38 |
| **Beta** | | | | |
| Participant Intercept | 199.51 | 14.12 | 9.27 | 18.62 |
| Residual | 40.97 | 6.40 | 3.76 | 9.12 |
| **Gamma** | | | | |
| Participant Intercept | 149.40 | 12.22 | 7.25 | 16.55 |
| Residual | 42.15 | 6.49 | 3.65 | 9.45 |

#### 3.2 Predictor: Aperiodic Exponent

##### 3.2.1 Unadjusted model

**Supplementary Table 8. In-detail fixed effect results for linear mixed-effect model analyses on dystonia severity.** Predictor: aperiodic exponent (average across LFP channels). Covariates: none (unadjusted analyses).

| Fixed effects | β | 95% CI for β (lower) | 95% CI for β (upper) | *SE* | df | t | *P* value *(uncorr.)* | *P* value *(FDR corrected)* |
|---|---|---|---|---|---|---|---|---|
| Intercept | 74.90 | 51.00 | 98.85 | 11.76 | 38.38 | 6.37 | <0.001 | |
| Aperiodic Exponent | 7.45 | -5.94 | 21.06 | 6.73 | 33.61 | 1.11 | 0.274 | n/a |

**Supplementary Table 9. In-detail random effect results for linear mixed-effect model analyses on dystonia severity.** Predictor: aperiodic exponent (average across LFP channels). Covariates: none (unadjusted analyses).

| Random effects | Variance | SD | 95% CI for SD (lower) | 95% CI for SD (upper) |
|---|---|---|---|---|
| Participant Intercept | 730.76 | 27.03 | 19.75 | 34.36 |
| Residual | 43.30 | 6.58 | 4.02 | 9.17 |

##### 3.2.2 Adjusted model

**Model specifications and diagnostics.**

When the aperiodic exponent was examined, fixed effects results indicated that this was not a significant predictor of dystonia severity ($P$ = 0.79; Table 2 and Supplementary Table 10). Sensitivity analyses accounting for heteroskedasticity ($\chi^2(1) = 0.93$, $P = 0.33$) did not change these conclusions (aperiodic exponent β = -1.39, SE = 4.82, $P$ = 0.77).

**Supplementary Table 10. In-detail fixed effect results for linear mixed-effect model analyses on dystonia severity.** Predictor: aperiodic exponent (average across LFP channels). Covariates: pallidal involvement and medication number.

| Fixed effect | β | 95% CI for β (lower) | 95% CI for β (upper) | *SE* | df | t | *P* value *(uncorr.)* | *P* value *(FDR corrected)* |
|---|---|---|---|---|---|---|---|---|
| Intercept | 41.81 | 23.86 | 59.73 | 8.97 | 33.68 | 4.66 | <0.001 | |
| Aperiodic Exponent | -1.25 | -11.28 | 9.17 | 5.00 | 36.80 | -0.25 | 0.790 | n/a |
| Pallidal involvement | 13.80 | 1.89 | 25.50 | 6.04 | 25.60 | 2.28 | 0.027 | |
| Medication number | n/a | n/a | n/a | n/a | 3 | $\chi^2 = 33.93^a$ | <0.001 | |
| [a] $\chi^2$ for medication number compares a model without the medication term to one including medication number modelled as a natural spline (with degrees of freedom equal to the number reported). | | | | | | | | |

**Supplementary Table 11. In-detail random effect results for linear mixed-effect model analyses on dystonia severity.** Predictor: aperiodic exponent (average across LFP channels). Covariates: pallidal involvement and medication number.

| Random effect | Variance | SD | 95% CI for SD (lower) | 95% CI for SD (upper) |
|---|---|---|---|---|
| Participant Intercept | 198.69 | 14.1 | 9.02 | 18.70 |
| Residual | 44.83 | 6.7 | 3.98 | 9.43 |

#### 3.3 Predictor: Pallidopallidal coherence

##### 3.3.1 Unadjusted models

**Supplementary Table 12. In-detail fixed effect results for linear mixed-effect model analyses on dystonia severity.** Predictor: Pallidopallidal Coherence (best LFP channel). Covariates: none (unadjusted analyses).

| Fixed effects | β | 95% CI for β (lower) | 95% CI for β (upper) | *SE* | df | t | *P* value *(uncorr.)* | *P* value *(FDR corrected)* |
|---|---|---|---|---|---|---|---|---|
| **Delta** | | | | | | | | |
| Intercept | 84.30 | 67.36 | 101.48 | 8.70 | 30.54 | 9.69 | <0.001 | |

| | | | | | | | | |
|---|---|---|---|---|---|---|---|---|
| Pallidopallidal Coherence | 4.97 | -24.76 | 35.02 | 14.87 | 16.01 | 0.33 | 0.741 | 0.741 |
| **Theta** | | | | | | | | |
| Intercept | 81.65 | 63.23 | 99.80 | 9.28 | 29.37 | 8.80 | <0.001 | |
| Pallidopallidal Coherence | 13.76 | -27.82 | 56.12 | 21.28 | 16.80 | 0.65 | 0.542 | *0.677* |
| **Alpha** | | | | | | | | |
| Intercept | 89.66 | 77.13 | 102.46 | 6.44 | 40.73 | 13.92 | <0.001 | |
| Pallidopallidal Coherence | -11.49 | -39.99 | 17.80 | 14.97 | 14.86 | -0.77 | 0.445 | *0.677* |
| **Beta** | | | | | | | | |
| Intercept | 95.90 | 83.96 | 107.80 | 6.11 | 40.86 | 15.69 | <0.001 | |
| Pallidopallidal Coherence | -53.02 | -94.27 | -12.43 | 20.95 | 14.40 | -2.53 | 0.012 | *0.060* |
| **Gamma** | | | | | | | | |
| Intercept | 88.67 | 77.91 | 99.16 | 5.44 | 35.47 | 16.31 | <0.001 | |
| Pallidopallidal Coherence | -23.20 | -65.54 | 18.76 | 21.46 | 12.49 | -1.08 | 0.268 | *0.067* |

**Supplementary Table 13. In-detail random effect results for linear mixed-effect model analyses on dystonia severity.** Predictor: Pallidopallidal Coherence (best LFP channel). Covariates: none (unadjusted analyses).

| **Random effects** | **Variance** | **SD** | **95% CI for SD (lower)** | **95% CI for SD (upper)** |
|---|---|---|---|---|
| **Delta** | | | | |
| Participant Intercept | 757.80 | 27.53 | 20.24 | 34.90 |
| Residual | 43.59 | 6.60 | 3.98 | 9.32 |
| **Theta** | | | | |
| Participant Intercept | 765.23 | 27.66 | 20.30 | 35.03 |
| Residual | 41.80 | 6.47 | 3.91 | 9.04 |
| **Alpha** | | | | |
| Participant Intercept | 751.22 | 27.41 | 20.07 | 34.77 |
| Residual | 42.84 | 6.55 | 3.99 | 9.16 |
| **Beta** | | | | |
| Participant Intercept | 693.24 | 26.33 | 19.42 | 33.37 |
| Residual | 34.09 | 5.84 | 3.54 | 8.19 |
| **Gamma** | | | | |
| Participant Intercept | 748.76 | 27.36 | 20.14 | 34.69 |
| Residual | 41.36 | 6.43 | 3.93 | 8.98 |

#### 3.3.2 Adjusted models

**Supplementary Table 14. In-detail fixed effect results for linear mixed-effect model analyses on dystonia severity.** Predictor: Pallidopallidal Coherence (best LFP channel). Covariates: pallidal involvement and medication number.

| **Fixed effect** | **β** | **95% CI for β (lower)** | **95% CI for β (upper)** | ***SE*** | **df** | **t** | ***P* value (*uncorr.*)** | ***P* value (*FDR corrected*)** |
|---|---|---|---|---|---|---|---|---|
| **Delta** | | | | | | | | |
| Intercept | 44.19 | 27.52 | 60.76 | 8.16 | 37.68 | 5.41 | <0.001 | |
| Pallidopallidal Coherence | -12.15 | -39.26 | 15.22 | 13.50 | 23.58 | -0.90 | 0.393 | 0.393 |
| Pallidal involvement | 14.09 | 2.86 | 25.15 | 5.62 | 25.74 | 2.51 | 0.016 | |
| Medication number | n/a | n/a | n/a | n/a | 2 | $\chi^2$ = 35.03 | <0.001 | |
| **Theta** | | | | | | | | |
| Intercept | 49.04 | 31.80 | 65.91 | 8.58 | 36.82 | 5.72 | <0.001 | |
| Pallidopallidal Coherence | -31.20 | -70.05 | 7.42 | 19.51 | 31.30 | -1.60 | 0.120 | 0.200 |
| Pallidal involvement | 14.43 | 3.60 | 25.08 | 5.44 | 23.11 | 2.65 | 0.012 | |
| Medication number | n/a | n/a | n/a | n/a | 3 | $\chi^2$ = 36.81[a] | <0.001 | |
| **Alpha** | | | | | | | | |
| Intercept | 46.00 | 32.29 | 59.75 | 6.90 | 33.91 | 6.67 | <0.001 | |
| Pallidopallidal Coherence | -27.79 | -55.51 | 0.18 | 13.92 | 22.91 | -2.00 | 0.051 | 0.127 |
| Pallidal involvement | 15.33 | 4.21 | 25.98 | 5.46 | 23.43 | 2.81 | 0.008 | |
| Medication number | n/a | n/a | n/a | n/a | 3 | $\chi^2$ = 37.29 | <0.001 | |
| **Beta** | | | | | | | | |
| Intercept | 52.24 | 39.45 | 65.33 | 6.69 | 36.08 | 7.81 | <0.001 | |

| | | | | | | | | |
|---|---|---|---|---|---|---|---|---|
| Pallidopallidal Coherence | -65.42 | -103.72 | -27.43 | 18.84 | 24.33 | -3.47 | 0.002 | 0.01 |
| Pallidal involvement | 14.15 | 4.13 | 23.80 | 4.99 | 22.80 | 2.84 | 0.006 | |
| Medication number | n/a | n/a | n/a | n/a | 3 | $\chi^2$ = 39.29 | <0.001 | |
| **Gamma** | | | | | | | | |
| Intercept | 41.86 | 29.03 | 54.75 | 6.38 | 33.88 | 6.57 | <0.001 | |
| Pallidopallidal Coherence | -25.14 | -67.89 | 17.98 | 21.87 | 14.97 | -1.15 | 0.253 | 0.316 |
| Pallidal involvement | 13.81 | 2.61 | 24.86 | 5.63 | 25.65 | 2.45 | 0.020 | |
| Medication number | n/a | n/a | n/a | n/a | 2 | $\chi^2$ = 34.12 | <0.001 | |
| [a] $\chi^2$ for medication number compares a model without the medication term to one including medication number modelled as a natural spline (with degrees of freedom equal to the number reported). | | | | | | | | |

**Supplementary Table 15. In-detail random effect results for linear mixed-effect model analyses on dystonia severity.** Predictor: Pallidopallidal Coherence (best LFP channel). Covariates: pallidal involvement and medication number.

| Random effect | Variance | SD | 95% CI for SD (lower) | 95% CI for SD (upper) |
|---|---|---|---|---|
| **Delta** | | | | |
| Participant Intercept | 172.37 | 13.13 | 8.19 | 17.49 |
| Residual | 49.14 | 7.01 | 4.18 | 9.92 |
| **Theta** | | | | |
| Participant Intercept | 155.69 | 12.48 | 7.34 | 16.87 |
| Residual | 53.95 | 7.35 | 4.40 | 10.35 |
| **Alpha** | | | | |
| Participant Intercept | 158.11 | 12.57 | 7.55 | 16.85 |
| Residual | 48.44 | 6.96 | 4.14 | 9.89 |
| **Beta** | | | | |
| Participant Intercept | 140.35 | 11.85 | 7.42 | 15.85 |
| Residual | 37.16 | 6.10 | 3.61 | 8.67 |
| **Gamma** | | | | |
| Participant Intercept | 180.93 | 13.45 | 8.78 | 17.81 |
| Residual | 44.54 | 6.67 | 4.00 | 9.42 |

##### 3.3.3 Adjusted models with interaction term

**Supplementary Table 16. In-detail fixed effect results for linear mixed-effect model analyses on dystonia severity.** Predictor: Pallidopallidal Coherence (best LFP channel). Covariates: pallidal involvement, medication number, and interaction term: pallidopallidal coherence x medication number (3 df).

| Fixed effect | β | 95% CI for β (lower) | 95% CI for β (upper) | *SE* | df | t | ***P* value (*uncorr.*)** | ***P* value (*FDR corrected*)** |
|---|---|---|---|---|---|---|---|---|
| **Delta** | | | | | | | | |
| Intercept | 107.29 | 68.43 | 145.66 | 18.95 | 35.51 | 5.66 | <0.001 | |
| Pallidopallidal Coherence | -164.31 | -249.97 | -77.65 | 42.85 | 34.94 | -3.83 | <0.001 | |
| Pallidal involvement | 13.90 | 5.00 | 22.88 | 4.55 | 22.55 | 3.05 | 0.002 | |
| Medication number | n/a | n/a | n/a | n/a | 2 | $\chi^2$ = 35.04 | <0.001 | |
| Coherence x Medication number | n/a | n/a | n/a | n/a | 2 | $\chi^2$ = 10.51 | *0.005* | *0.005* |
| **Theta** | | | | | | | | |
| Intercept | 132.57 | 90.06 | 174.97 | 21.71 | 29.56 | 6.11 | <0.001 | |
| Pallidopallidal Coherence | -302.02 | -437.91 | -168.12 | 68.10 | 28.76 | -4.43 | <0.001 | |
| Pallidal involvement | 10.98 | 2.60 | 19.41 | 4.26 | 22.39 | 2.58 | 0.010 | |
| Medication number | n/a | n/a | n/a | n/a | 3 | $\chi^2$ = 36.81 | <0.001 | |
| Coherence x Medication number | n/a | n/a | n/a | n/a | 3 | $\chi^2$ = 17.02 | <0.001 | <0.001 |
| **Alpha** | | | | | | | | |
| Intercept | 99.16 | 75.11 | 123.97 | 12.32 | 33.05 | 8.05 | <0.001 | |
| Pallidopallidal Coherence | -273.34 | -381.79 | -166.39 | 54.25 | 32.67 | -5.04 | <0.001 | |
| Pallidal involvement | 15.88 | 7.80 | 23.92 | 4.06 | 24.65 | 3.91 | <0.001 | |
| Medication number | n/a | n/a | n/a | n/a | 3 | $\chi^2$ = 37.29 | <0.001 | |

| | | | | | | | | |
|---|---|---|---|---|---|---|---|---|
| Coherence x Medication number | n/a | n/a | n/a | n/a | 3 | χ² = 27.42 | <0.001 | <0.001 |
| **Beta** | | | | | | | | |
| Intercept | 96.06 | 76.21 | 116.11 | 9.84 | 31.99 | 9.77 | <0.001 | |
| Pallidopallidal Coherence | -308.50 | -413.43 | -203.53 | 51.00 | 33.03 | -6.05 | <0.001 | |
| Pallidal involvement | 13.21 | 4.86 | 21.11 | 4.06 | 25.92 | 3.25 | 0.001 | |
| Medication number | n/a | n/a | n/a | n/a | 3 | χ² = 39.29 | <0.001 | |
| Coherence x Medication number | n/a | n/a | n/a | n/a | 3 | χ² = 32.96 | <0.001 | <0.001 |
| **Gamma** | | | | | | | | |
| Intercept | 93.12 | 73.30 | 112.33 | 9.83 | 34.99 | 9.47 | <0.001 | |
| Pallidopallidal Coherence | -608.88 | -804.04 | -404.17 | 99.10 | 34.42 | -6.14 | <0.001 | |
| Pallidal involvement | 12.90 | 5.98 | 19.94 | 3.53 | 22.66 | 3.65 | <0.001 | |
| Medication number | n/a | n/a | n/a | n/a | 2 | χ² = 34.12 | <0.001 | |
| Coherence x Medication number | n/a | n/a | n/a | n/a | 2 | χ² = 26.17 | <0.001 | <0.001 |

[a] χ² for medication number compares a model without the medication term to one including medication number modelled as a natural spline (with degrees of freedom equal to the number reported).

**Supplementary Table 17. In-detail random effect results for linear mixed-effect model analyses on dystonia severity.** Predictor: Pallidopallidal Coherence (best LFP channel). Covariates: pallidal involvement, medication number, and interaction term: pallidopallidal coherence x medication number (3 df).

| Random effect | Variance | SD | 95% CI for SD (lower) | 95% CI for SD (upper) |
|---|---|---|---|---|
| **Delta** | | | | |
| Participant Intercept | 69.47 | 8.33 | 0.00 | 12.97 |
| Residual | 79.67 | 8.93 | 5.31 | 12.44 |
| **Theta** | | | | |
| Participant Intercept | 50.58 | 7.11 | 0.00 | 11.77 |
| Residual | 76.74 | 8.76 | 5.16 | 11.93 |
| **Alpha** | | | | |
| Participant Intercept | 77.98 | 8.83 | 3.89 | 12.27 |
| Residual | 33.44 | 5.78 | 3.23 | 8.42 |
| **Beta** | | | | |
| Participant Intercept | 102.18 | 10.11 | 6.86 | 13.21 |
| Residual | 10.43 | 3.23 | 1.73 | 4.75 |
| **Gamma** | | | | |
| Participant Intercept | 39.38 | 6.28 | 0.00 | 10.14 |
| Residual | 60.38 | 7.77 | 4.47 | 10.63 |

### 3.4 Control analyses. Predictor: Peak power (best LFP channel) of 3 Hz narrow band around the peak

#### 3.4.1 Adjusted models

**Supplementary Table 18. In-detail fixed effect results for linear mixed-effect model analyses on dystonia severity.** Predictor: power of a 3 Hz narrow band around the peak (best LFP channel). Covariates: pallidal involvement and medication number.

| Fixed effect | β | 95% CI for β (lower) | 95% CI for β (upper) | *SE* | df | t | *P* value (uncorr.) | *P* value *(FDR corrected)* |
|---|---|---|---|---|---|---|---|---|
| **Delta** | | | | | | | | |
| Intercept | 45.87 | 33.26 | 58.96 | 6.63 | 32.17 | 6.92 | <0.001 | |
| Peak power | -5.05 | -9.11 | -0.82 | 2.01 | 16.07 | -2.51 | 0.016 | 0.062 |
| Pallidal involvement | 15.09 | 3.68 | 26.09 | 5.66 | 25.30 | 2.66 | 0.010 | |
| Medication number | n/a | n/a | n/a | n/a | 3 | χ² = 29.06 [a] | <0.001 | |
| **Theta** | | | | | | | | |
| Intercept | 43.82 | 29.55 | 58.03 | 7.19 | 29.90 | 6.09 | <0.001 | |

| | | | | | | | | |
|---|---|---|---|---|---|---|---|---|
| Peak power | -4.39 | -11.85 | 3.66 | 3.79 | 34.22 | -1.16 | 0.273 | 0.296 |
| Pallidal involvement | 12.30 | 0.18 | 23.97 | 5.96 | 24.70 | 2.06 | 0.047 | |
| Medication number | n/a | n/a | n/a | n/a | 3 | $\chi^2$ = 31.77 [a] | <0.001 | |
| **Alpha** | | | | | | | | |
| Intercept | 41.52 | 28.28 | 54.71 | 6.65 | 30.14 | 6.24 | <0.001 | |
| Peak power | -16.50 | -46.16 | 14.76 | 15.11 | 27.10 | -1.09 | 0.296 | 0.296 |
| Pallidal involvement | 14.64 | 2.59 | 26.11 | 5.90 | 25.85 | 2.48 | 0.019 | |
| Medication number | n/a | n/a | n/a | n/a | 3 | $\chi^2$ = 34.32 [a] | <0.001 | |
| **Beta** | | | | | | | | |
| Intercept | 41.52 | 28.32 | 54.67 | 6.64 | 31.05 | 6.25 | <0.001 | |
| Peak power | -19.83 | -50.89 | 12.49 | 15.77 | 14.50 | -1.26 | 0.225 | 0.296 |
| Pallidal involvement | 14.13 | 2.22 | 25.58 | 5.86 | 26.02 | 2.41 | 0.022 | |
| Medication number | n/a | n/a | n/a | n/a | 3 | $\chi^2$ = 33.60 [a] | <0.001 | |
| **Gamma** | | | | | | | | |
| Intercept | 51.28 | 36.13 | 66.11 | 7.59 | 34.18 | 6.76 | <0.001 | |
| Peak power | n/a | n/a | n/a | n/a | 2 | $\chi^2$ = 7.32 [b] | *0.025* | 0.062 |
| Pallidal involvement | 10.68 | 0.17 | 20.89 | 5.23 | 24.76 | 2.04 | 0.048 | |
| Medication number | n/a | n/a | n/a | n/a | 3 | $\chi^2$ = 34.09 [a] | <0.001 | |

[a] $\chi^2$ for medication number compares a model without the medication term to one including medication number modelled as a natural spline (with degrees of freedom equal to the number reported).
[b] $\chi^2$ for gamma peak power compares a model without peak power to one with peak power modelled as a natural spline (2 df). The relationship between peak power and BFMDRS-M was modelled using a natural spline (2 df) as this significantly improved both model fit ($\chi^2(1) = 9.45$, ***P*** = 0.002) and the distribution of residuals relative to a linear term

**Supplementary Table 19. In-detail random effect results for linear mixed-effect model analyses on dystonia severity.** Predictor: power of a 3 Hz narrow band around the peak (best LFP channel). Covariates: pallidal involvement and medication number.

| Random effect | Variance | SD | 95% CI for SD (lower) | 95% CI for SD (upper) |
|---|---|---|---|---|
| **Delta** | | | | |
| Participant Intercept | 190.57 | 13.80 | 9.20 | 18.15 |
| Residual | 33.09 | 5.75 | 3.36 | 8.19 |
| **Theta** | | | | |
| Participant Intercept | 203.56 | 14.27 | 9.28 | 18.84 |
| Residual | 40.13 | 6.33 | 3.78 | 8.94 |
| **Alpha** | | | | |
| Participant Intercept | 196.43 | 14.02 | 9.16 | 18.47 |
| Residual | 42.43 | 6.51 | 3.87 | 9.27 |
| **Beta** | | | | |
| Participant Intercept | 201.23 | 14.19 | 9.37 | 18.67 |
| Residual | 40.15 | 6.34 | 3.71 | 9.00 |
| **Gamma** | | | | |
| Participant Intercept | 140.08 | 11.84 | 6.64 | 16.09 |
| Residual | 51.29 | 7.16 | 4.11 | 10.14 |

### 3.5 Control analyses. Predictor: Peak power (average across all LFP channels)

#### 3.5.1 Adjusted models

**Supplementary Table 20. In-detail fixed effect results for linear mixed-effect model analyses on dystonia severity.** Predictor: peak power (average across all LFP channels). Covariates: pallidal involvement and medication number.

| Fixed effect | β | 95% CI for β (lower) | 95% CI for β (upper) | *SE* | df | t | ***P*** **value (uncorr.)** | ***P*** **value (FDR corrected)** |
|---|---|---|---|---|---|---|---|---|
| **Delta** | | | | | | | | |
| Intercept | 46.55 | 32.80 | 60.80 | 7.22 | 29.24 | 6.45 | <0.001 | |
| Peak power | -5.37 | -9.13 | -1.48 | 1.89 | 17.97 | -2.84 | 0.007 | *0.035* |
| Pallidal involvement | 12.13 | -0.21 | 24.17 | 6.16 | 23.98 | 1.97 | 0.055 | |
| Medication number | n/a | n/a | n/a | n/a | 4 | $\chi^2$ = 30.32 [a] | <0.001 | |
| **Theta** | | | | | | | | |

| | | | | | | | | |
|---|---|---|---|---|---|---|---|---|
| Intercept | 41.92 | 28.53 | 55.46 | 6.88 | 30.62 | 6.09 | <0.001 | |
| Peak power | -4.29 | -15.33 | 6.88 | 5.61 | 37.08 | -0.76 | 0.460 | 0.693 |
| Pallidal involvement | 13.25 | 1.79 | 24.42 | 5.69 | 25.22 | 2.33 | 0.026 | |
| Medication number | n/a | n/a | n/a | n/a | 2 | $\chi^2$ = 33.56 [a] | <0.001 | |
| **Alpha** | | | | | | | | |
| Intercept | 41.32 | 27.41 | 54.82 | 6.91 | 30.31 | 5.98 | <0.001 | |
| Peak power | -10.41 | -50.33 | 30.92 | 20.07 | 31.17 | -0.52 | 0.635 | 0.693 |
| Pallidal involvement | 14.07 | 1.99 | 25.90 | 5.96 | 25.81 | 2.36 | 0.024 | |
| Medication number | n/a | n/a | n/a | n/a | 3 | $\chi^2$ = 32.87 [a] | <0.001 | |
| **Beta** | | | | | | | | |
| Intercept | 40.58 | 27.18 | 53.87 | 6.69 | 29.29 | 6.07 | <0.001 | |
| Peak power | -4.97 | -29.02 | 19.56 | 12.03 | 33.80 | -0.41 | 0.693 | 0.693 |
| Pallidal involvement | 13.92 | 2.03 | 25.52 | 6.00 | 24.93 | 2.32 | 0.027 | |
| Medication number | n/a | n/a | n/a | n/a | 3 | $\chi^2$ = 34.14 [a] | <0.001 | |
| **Gamma** | | | | | | | | |
| Intercept | 44.74 | 26.96 | 62.34 | 9.04 | 28.34 | 4.95 | <0.001 | |
| Peak power | n/a | n/a | n/a | n/a | 2 | $\chi^2$ = 1.01 [b] | *0.600* | 0.693 |
| Pallidal involvement | 12.74 | 0.82 | 24.16 | 5.86 | 24.19 | 2.18 | 0.036 | |
| Medication number | n/a | n/a | n/a | n/a | 3 | $\chi^2$ = 29.47 [a] | <0.001 | |

[a] $\chi^2$ for medication number compares a model without the medication term to one including medication number modelled as a natural spline (with degrees of freedom equal to the number reported).
[b] $\chi^2$ for gamma peak power compares a model without peak power to one with peak power modelled as a natural spline (2 df). The relationship between peak power and BFMDRS-M was modelled using a natural spline (2 df) as this significantly improved both model fit ($\chi^2(1) = 9.45$, ***P*** = 0.002) and the distribution of residuals relative to a linear term

**Supplementary Table 21. In-detail random effect results for linear mixed-effect model analyses on dystonia severity.** Predictor: peak power (average across all LFP channels). Covariates: pallidal involvement and medication number.

| **Random effect** | **Variance** | **SD** | **95% CI for SD (lower)** | **95% CI for SD (upper)** |
|---|---|---|---|---|
| **Delta** | | | | |
| Participant Intercept | 225.49 | 15.02 | 10.28 | 19.70 |
| Residual | 26.29 | 5.13 | 2.98 | 7.33 |
| **Theta** | | | | |
| Participant Intercept | 186.55 | 13.66 | 8.84 | 18.05 |
| Residual | 44.68 | 6.68 | 4.04 | 9.40 |
| **Alpha** | | | | |
| Participant Intercept | 197.10 | 14.04 | 9.18 | 18.55 |
| Residual | 44.73 | 6.69 | 3.99 | 9.46 |
| **Beta** | | | | |
| Participant Intercept | 203.79 | 14.28 | 9.26 | 18.94 |
| Residual | 43.04 | 6.56 | 3.97 | 9.26 |
| **Gamma** | | | | |
| Participant Intercept | 184.38 | 13.58 | 8.33 | 18.23 |
| Residual | 50.42 | 7.10 | 4.13 | 10.07 |

### 3.6 Control analyses. Predictor: Pallidopallidal coherence (average across all LFP channels).

#### 3.6.1 Adjusted models with interaction term

**Supplementary Table 22. In-detail fixed effect results for linear mixed-effect model analyses on dystonia severity.** Pallidopallidal Coherence (best LFP channel). Covariates: pallidal involvement, medication number, and interaction term: pallidopallidal coherence x medication number (3 df).

| **Fixed effect** | **β** | **95% CI for β (lower)** | **95% CI for β (upper)** | ***SE*** | **df** | **t** | ***P* value (*uncorr.*)** | ***P* value (*FDR corrected*)** |
|---|---|---|---|---|---|---|---|---|
| **Delta** | | | | | | | | |
| Intercept | 79.09 | 47.35 | 111.41 | 15.55 | 36.00 | 5.09 | <0.001 | |
| Pallidopallidal Coherence | -174.79 | -299.31 | -49.88 | 61.05 | 35.91 | -2.86 | 0.010 | |
| Pallidal involvement | 14.92 | 5.51 | 24.62 | 4.90 | 19.59 | 3.05 | 0.003 | |

| | | | | | | | | |
|---|---|---|---|---|---|---|---|---|
| Medication number | n/a | n/a | n/a | n/a | 2 | $\chi^2$ = 33.38 | <0.001 | |
| Coherence x Medication number | n/a | n/a | n/a | n/a | 2 | $\chi^2$ =5.66 | *0.058* | *0.058* |
| **Theta** | | | | | | | | |
| Intercept | 102.73 | 64.77 | 141.63 | 19.49 | 27.49 | 5.27 | <0.001 | |
| Pallidopallidal Coherence | -365.84 | -580.29 | -153.90 | 106.35 | 26.35 | -3.44 | 0.001 | |
| Pallidal involvement | 13.53 | 4.31 | 22.68 | 4.69 | 21.94 | 2.88 | 0.005 | |
| Medication number | n/a | n/a | n/a | n/a | 3 | $\chi^2$ = 34.96 | <0.001 | |
| Coherence x Medication number | n/a | n/a | n/a | n/a | 3 | $\chi^2$ = 11.46 | *0.009* | 0.011 |
| **Alpha** | | | | | | | | |
| Intercept | 106.49 | 85.39 | 127.05 | 10.24 | 30.73 | 10.40 | <0.001 | |
| Pallidopallidal Coherence | -568.76 | -738.93 | -389.17 | 84.46 | 30.18 | -6.73 | <0.001 | |
| Pallidal involvement | 18.53 | 10.34 | 26.51 | 4.08 | 26.69 | 4.54 | <0.001 | |
| Medication number | n/a | n/a | n/a | n/a | 3 | $\chi^2$ = 37.56 | <0.001 | |
| Coherence x Medication number | n/a | n/a | n/a | n/a | 3 | $\chi^2$ = 40.55 | <0.001 | <0.001 |
| **Beta** | | | | | | | | |
| Intercept | 86.63 | 60.59 | 111.66 | 12.75 | 33.36 | 6.80 | <0.001 | |
| Pallidopallidal Coherence | -476.87 | -733.24 | -209.78 | 129.33 | 32.11 | -3.69 | <0.001 | |
| Pallidal involvement | 14.10 | 4.60 | 23.16 | 4.71 | 25.18 | 2.99 | 0.003 | |
| Medication number | n/a | n/a | n/a | n/a | 3 | $\chi^2$ = 39.80 | <0.001 | |
| Coherence x Medication number | n/a | n/a | n/a | n/a | 3 | $\chi^2$ = 18.88 | <0.001 | <0.001 |
| **Gamma** | | | | | | | | |
| Intercept | 93.81 | 72.37 | 115.68 | 10.65 | 35.26 | 8.80 | <0.001 | |
| Pallidopallidal Coherence | -954.65 | -1300.31 | -599.37 | 172.19 | 34.73 | -5.54 | <0.001 | |
| Pallidal involvement | 12.32 | 3.71 | 20.72 | 4.27 | 25.99 | 2.89 | 0.005 | |
| Medication number | n/a | n/a | n/a | n/a | 2 | $\chi^2$ = 34.01 | <0.001 | |
| Coherence x Medication number | n/a | n/a | n/a | n/a | 2 | $\chi^2$ = 27.37 | <0.001 | <0.001 |
| [a] $\chi^2$ for medication number compares a model without the medication term to one including medication number modelled as a natural spline (with degrees of freedom equal to the number reported). | | | | | | | | |

**Supplementary Table 23. In-detail random effect results for linear mixed-effect model analyses on dystonia severity.** Pallidopallidal Coherence (best LFP channel). Covariates: pallidal involvement, medication number, and interaction term: pallidopallidal coherence x medication number (3 df).

| **Random effect** | **Variance** | **SD** | **95% CI for SD (lower)** | **95% CI for SD (upper)** |
|---|---|---|---|---|
| **Delta** | | | | |
| Participant Intercept | 88.31 | 9.40 | 0.00 | 14.16 |
| Residual | 83.44 | 9.13 | 5.35 | 12.79 |
| **Theta** | | | | |
| Participant Intercept | 81.45 | 9.03 | 0.00 | 13.66 |
| Residual | 74.08 | 8.61 | 4.99 | 12.06 |
| **Alpha** | | | | |
| Participant Intercept | 97.47 | 9.87 | 6.41 | 12.92 |
| Residual | 11.45 | 3.38 | 1.76 | 5.00 |
| **Beta** | | | | |
| Participant Intercept | 136.70 | 11.69 | 7.88 | 15.26 |
| Residual | 13.27 | 3.64 | 1.95 | 5.37 |
| **Gamma** | | | | |
| Participant Intercept | 105.59 | 10.28 | 6.73 | 13.61 |
| Residual | 23.41 | 4.84 | 2.71 | 6.98 |

### 3.7 Control analyses controlling for the potentially confounding effect of dystonia aetiology.

#### 3.7.1 Adjusted models

**Supplementary Table 24. In-detail fixed effect results for linear mixed-effect model analyses on dystonia severity, when controlling for the potentially confounding effect of dystonia aetiology.** Predictors are specified in row headers. Covariates: pallidal involvement, medication number, dystonia aetiology, and for pallidopallidal coherence, interaction term: pallidopallidal coherence x medication number (3 df).

| Fixed effect | β | 95% CI for β (lower) | 95% CI for β (upper) | *SE* | df | t | *P* value (*uncorr.)* |
|---|---|---|---|---|---|---|---|
| **Delta Peak Power (best LFP channel)** | | | | | | | |
| Intercept | 44.23 | 31.31 | 57.64 | 6.68 | 30.27 | 6.62 | <0.001 |
| Delta Peak Power | -2.57 | -4.74 | -0.33 | 1.08 | 13.78 | -2.38 | **0.022** |
| Pallidal involvement | 13.39 | 0.84 | 24.95 | 6.03 | 24.25 | 2.22 | 0.034 |
| Medication number | - | - | - | - | 3 | $\chi^2$ = 22.11 | <0.001 |
| Dystonia aetiology | 5.64 | -9.45 | 21.34 | 7.89 | 27.17 | 0.71 | 0.470 |
| **Gamma Peak Power (best LFP channel)** | | | | | | | |
| Intercept | 52.79 | 36.28 | 68.81 | 8.31 | 34.98 | 6.35 | <0.001 |
| Gamma Peak Power | - | - | - | - | 2 | $\chi^2$ = 8.18 | **0.016** |
| Pallidal involvement | 10.60 | -1.05 | 21.68 | 5.68 | 24.47 | 1.87 | 0.068 |
| Medication number | - | - | - | - | 2 | $\chi^2$ = 29.48 | <0.001 |
| Dystonia aetiology | -3.75 | -20.62 | 13.73 | 8.69 | 31.61 | -0.43 | 0.686 |
| **Delta Pallidopallidal coherence (best LFP channel)** | | | | | | | |
| Intercept | 104.70 | 65.06 | 145.15 | 19.57 | 34.71 | 5.35 | <0.001 |
| Pallidopallidal Coherence | -158.24 | -246.23 | -69.00 | 44.29 | 34.42 | -3.57 | <0.001 |
| Pallidal involvement | 13.37 | 3.76 | 23.00 | 4.86 | 21.63 | 2.75 | 0.006 |
| Medication number | - | - | - | - | 2 | $\chi^2$ =25.39 | <0.001 |
| Coherence x Medication number | - | - | - | - | 2 | $\chi^2$ = 9.82 | **0.007** |
| Dystonia aetiology | 2.22 | -10.10 | 14.59 | 6.38 | 21.84 | 0.35 | 0.724 |
| **Theta Pallidopallidal coherence (best LFP channel)** | | | | | | | |
| Intercept | 138.94 | 92.82 | 186.09 | 23.54 | 29.53 | 5.90 | <0.001 |
| Pallidopallidal Coherence | -321.39 | -468.50 | -176.97 | 73.54 | 29.06 | -4.37 | <0.001 |
| Pallidal involvement | 11.75 | 2.98 | 20.61 | 4.42 | 21.30 | 2.66 | 0.008 |
| Medication number | - | - | - | - | 3 | $\chi^2$ = 27.05 | <0.001 |
| Coherence x Medication number | - | - | - | - | 3 | $\chi^2$ = 17.25 | **<0.001** |
| Dystonia aetiology | -4.51 | -16.51 | 7.35 | 6.19 | 23.40 | -0.73 | 0.483 |
| **Alpha Pallidopallidal coherence (best LFP channel)** | | | | | | | |
| Intercept | 100.03 | 74.84 | 125.49 | 12.70 | 31.85 | 7.88 | <0.001 |
| Pallidopallidal Coherence | -276.36 | -387.33 | -165.69 | 55.56 | 31.54 | -4.97 | <0.001 |
| Pallidal involvement | 16.44 | 7.66 | 25.05 | 4.38 | 23.25 | 3.75 | <0.001 |
| Medication number | - | - | - | - | 3 | $\chi^2$ = 27.38 | <0.001 |
| Coherence x Medication number | - | - | - | - | 3 | $\chi^2$ = 27.34 | **<0.001** |
| Dystonia aetiology | -2.14 | -12.98 | 9.30 | 5.77 | 24.33 | -0.37 | 0.724 |
| **Beta Pallidopallidal coherence (best LFP channel)** | | | | | | | |
| Intercept | 96.82 | 75.62 | 117.55 | 10.29 | 32.21 | 9.41 | <0.001 |
| Pallidopallidal Coherence | -311.65 | -421.76 | -201.14 | 52.74 | 32.57 | -5.91 | <0.001 |
| Pallidal involvement | 13.59 | 4.62 | 22.01 | 4.35 | 24.97 | 3.12 | <0.001 |
| Medication number | - | - | - | - | 3 | $\chi^2$ = 29.41 | <0.001 |
| Coherence x Medication number | - | - | - | - | 3 | $\chi^2$ = 32.84 | **<0.001** |
| Dystonia aetiology | -1.56 | -12.41 | 9.77 | 5.70 | 26.60 | -0.27 | 0.795 |
| **Gamma Pallidopallidal coherence (best LFP channel)** | | | | | | | |
| Intercept | 103.29 | 79.70 | 127.64 | 12.15 | 32.76 | 8.50 | <0.001 |
| Pallidopallidal Coherence | -734.09 | -994.14 | -475.53 | 129.98 | 32.70 | -5.65 | <0.001 |
| Pallidal involvement | 11.93 | 3.79 | 20.05 | 4.06 | 23.36 | 2.94 | 0.003 |
| Medication number | - | - | - | - | 3 | $\chi^2$ = 24.57 | <0.001 |
| Coherence x Medication number | - | - | - | - | 3 | $\chi^2$ = 30.33 | **<0.001** |
| Dystonia aetiology | 1.50 | -9.14 | 12.26 | 5.57 | 26.60 | 0.27 | 0.772 |

**Supplementary Table 25. In-detail random effect results for linear mixed-effect model analyses on dystonia severity, when controlling for the potentially confounding effect of dystonia aetiology.** Predictors are specified in row headers. Covariates: pallidal involvement, medication number, dystonia aetiology, and for pallidopallidal coherence, interaction term: pallidopallidal coherence x medication number (3 df).

| Random effect | Variance | SD | 95% CI for SD (lower) | 95% CI for SD (upper) |
|---|---|---|---|---|
| **Delta Peak Power (best LFP channel)** | | | | |
| Participant Intercept | 194.82 | 13.96 | 9.2 | 18.47 |

| | | | | |
|---|---|---|---|---|
| Residual | 33.77 | 5.81 | 3.4 | 8.27 |
| **Gamma Peak Power (best LFP channel)** | | | | |
| Participant Intercept | 162.04 | 12.73 | 7.78 | 17.06 |
| Residual | 41.56 | 6.45 | 3.68 | 9.19 |
| **Delta Pallidopallidal coherence (best LFP channel)** | | | | |
| Participant Intercept | 77.02 | 8.78 | 0.00 | 13.43 |
| Residual | 78.02 | 8.83 | 5.24 | 12.38 |
| **Theta Pallidopallidal coherence (best LFP channel)** | | | | |
| Participant Intercept | 51.33 | 7.16 | 0.00 | 11.91 |
| Residual | 77.78 | 8.82 | 5.23 | 12.03 |
| **Alpha Pallidopallidal coherence (best LFP channel)** | | | | |
| Participant Intercept | 80.15 | 8.95 | 3.79 | 12.47 |
| Residual | 34.24 | 5.85 | 3.26 | 8.56 |
| **Beta Pallidopallidal coherence (best LFP channel)** | | | | |
| Participant Intercept | 106.10 | 10.30 | 6.92 | 13.51 |
| Residual | 10.51 | 3.24 | 1.72 | 4.78 |
| **Gamma Pallidopallidal coherence (best LFP channel)** | | | | |
| Participant Intercept | 67.16 | 8.20 | 2.37 | 11.68 |
| Residual | 40.26 | 6.34 | 3.48 | 9.20 |

### 3.8 Sensitivity analyses excluding four recordings (*n* = 2 acquired during sleep, *n* = 2 for a participant with detected epileptiform discharges)

#### 3.8.1 Adjusted models

**Supplementary Table 26. In-detail fixed effect results for linear mixed-effect model sensitivity analyses on dystonia severity, excluding four Indefinite Streaming recordings.** Predictors are specified in row headers. Covariates: pallidal involvement, medication number, dystonia aetiology, and for pallidopallidal coherence, interaction term: pallidopallidal coherence x medication number (3 df).

| Fixed effect | β | 95% CI for β (lower) | 95% CI for β (upper) | *SE* | df | t | *P* value (*uncorr.)* |
|---|---|---|---|---|---|---|---|
| **Delta Peak Power (best LFP channel)** | | | | | | | |
| Intercept | 45.48 | 31.82 | 59.13 | 6.86 | 29.34 | 6.63 | <0.001 |
| Delta Peak Power | -2.89 | -5.37 | -0.39 | 1.21 | 12.32 | -2.38 | **0.024** |
| Pallidal involvement | 14.26 | 2.87 | 25.86 | 5.99 | 23.10 | 2.38 | 0.016 |
| Medication number | - | - | - | - | 3 | $\chi^2$ = 27.39 | <0.001 |
| **Gamma Peak Power (best LFP channel)** | | | | | | | |
| Intercept | 51.22 | 35.40 | 67.13 | 7.88 | 31.70 | 6.50 | <0.001 |
| Gamma Peak Power | - | - | - | - | 2 | $\chi^2$ = 7.51 | **0.023** |
| Pallidal involvement | 9.76 | -1.53 | 20.91 | 5.79 | 23.97 | 1.68 | 0.084 |
| Medication number | - | - | - | - | 3 | $\chi^2$ = 31.81 | <0.001 |
| **Delta Pallidopallidal coherence (best LFP channel)** | | | | | | | |
| Intercept | 109.07 | 68.11 | 151.48 | 20.43 | 31.73 | 5.34 | <0.001 |
| Pallidopallidal coherence | -168.81 | -263.74 | -77.12 | 46.44 | 31.23 | -3.64 | <0.001 |
| Pallidal involvement | 14.08 | 4.61 | 23.38 | 4.78 | 19.81 | 2.95 | 0.003 |
| Medication number | - | - | - | - | 2 | $\chi^2$ = 32.87 | <0.001 |
| Coherence x Medication number | - | - | - | - | 2 | $\chi^2$ = 9.48 | **0.008** |
| **Theta Pallidopallidal coherence (best LFP channel)** | | | | | | | |
| Intercept | 134.49 | 86.85 | 183.80 | 24.00 | 27.38 | 5.60 | <0.001 |
| Pallidopallidal coherence | -308.49 | -463.73 | -157.32 | 76.28 | 27.09 | -4.04 | <0.001 |
| Pallidal involvement | 11.45 | 2.96 | 19.91 | 4.36 | 18.70 | 2.63 | 0.010 |
| Medication number | - | - | - | - | 3 | $\chi^2$ = 36.38 | <0.001 |
| Coherence x Medication number | - | - | - | - | 3 | $\chi^2$ = 14.20 | **0.002** |
| **Alpha Pallidopallidal coherence (best LFP channel)** | | | | | | | |
| Intercept | 98.89 | 71.52 | 126.75 | 13.70 | 29.43 | 7.22 | <0.001 |
| Pallidopallidal coherence | -272.04 | -397.74 | -149.04 | 61.62 | 29.06 | -4.41 | <0.001 |
| Pallidal involvement | 15.83 | 7.36 | 24.22 | 4.31 | 21.48 | 3.67 | <0.001 |
| Medication number | - | - | - | - | 3 | $\chi^2$ = 34.68 | <0.001 |
| Coherence x Medication number | - | - | - | - | 3 | $\chi^2$ = 23.13 | **<0.001** |
| **Beta Pallidopallidal coherence (best LFP channel)** | | | | | | | |
| Intercept | 97.29 | 76.88 | 117.52 | 9.74 | 24.79 | 9.99 | <0.001 |
| Pallidopallidal coherence | -313.73 | -417.42 | -210.23 | 50.81 | 26.47 | -6.18 | <0.001 |
| Pallidal involvement | 12.56 | 4.21 | 21.03 | 4.35 | 23.83 | 2.89 | 0.004 |
| Medication number | - | - | - | - | 3 | $\chi^2$ = 36.27 | <0.001 |
| Coherence x Medication number | - | - | - | - | 3 | $\chi^2$ = 32.31 | **<0.001** |

| | | | | | | | |
|---|---|---|---|---|---|---|---|
| **Gamma Pallidopallidal coherence (best LFP channel)** | | | | | | | |
| Intercept | 94.25 | 73.24 | 115.30 | 10.46 | 30.99 | 9.01 | <0.001 |
| Pallidopallidal coherence | -621.03 | -839.66 | -414.02 | 105.52 | 30.67 | -5.89 | <0.001 |
| Pallidal involvement | 12.84 | 5.41 | 20.25 | 3.77 | 19.92 | 3.41 | 0.002 |
| Medication number | - | - | - | - | 2 | $\chi^2$ = 30.99 | <0.001 |
| Coherence x Medication number | - | - | - | - | 2 | $\chi^2$ = 23.96 | **<0.001** |

**Supplementary Table 27. In-detail random effect results for linear mixed-effect model sensitivity analyses on dystonia severity, excluding four Indefinite Streaming recordings.** Predictors are specified in row headers. Covariates: pallidal involvement, medication number, dystonia aetiology, and for pallidopallidal coherence, interaction term: pallidopallidal coherence x medication number (3 df).

| **Random effect** | **Variance** | **SD** | **95% CI for SD (lower)** | **95% CI for SD (upper)** |
|---|---|---|---|---|
| **Delta Peak Power (best LFP channel)** | | | | |
| Participant Intercept | 203.93 | 14.28 | 9.21 | 19.07 |
| Residual | 37.48 | 6.12 | 3.28 | 8.93 |
| **Gamma Peak Power (best LFP channel)** | | | | |
| Participant Intercept | 173.10 | 13.16 | 7.6 | 17.92 |
| Residual | 46.98 | 6.85 | 3.6 | 10.15 |
| **Delta Pallidopallidal coherence (best LFP channel)** | | | | |
| Participant Intercept | 67.89 | 8.24 | 0.00 | 13.51 |
| Residual | 96.33 | 9.81 | 5.26 | 13.49 |
| **Theta Pallidopallidal coherence (best LFP channel)** | | | | |
| Participant Intercept | 40.59 | 6.37 | 0.00 | 12.01 |
| Residual | 95.50 | 9.77 | 5.13 | 13.17 |
| **Alpha Pallidopallidal coherence (best LFP channel)** | | | | |
| Participant Intercept | 86.08 | 9.28 | 2.96 | 13.22 |
| Residual | 38.49 | 6.20 | 3.12 | 9.24 |
| **Beta Pallidopallidal coherence (best LFP channel)** | | | | |
| Participant Intercept | 115.42 | 10.74 | 7.18 | 14.21 |
| Residual | 8.59 | 2.93 | 1.29 | 4.56 |
| **Gamma Pallidopallidal coherence (best LFP channel)** | | | | |
| Participant Intercept | 38.02 | 6.17 | 0.00 | 10.79 |
| Residual | 72.42 | 8.51 | 4.42 | 11.53 |

### 3.9 Figures

**Supplementary Figure 2. Model-predicted association between BFMDRS-M scores and A) pallidal involvement and B) medication burden.** Graphs are based on the model using theta peak power (best LFP channel) as a predictor, but the estimated shape of the association is consistent across all models.

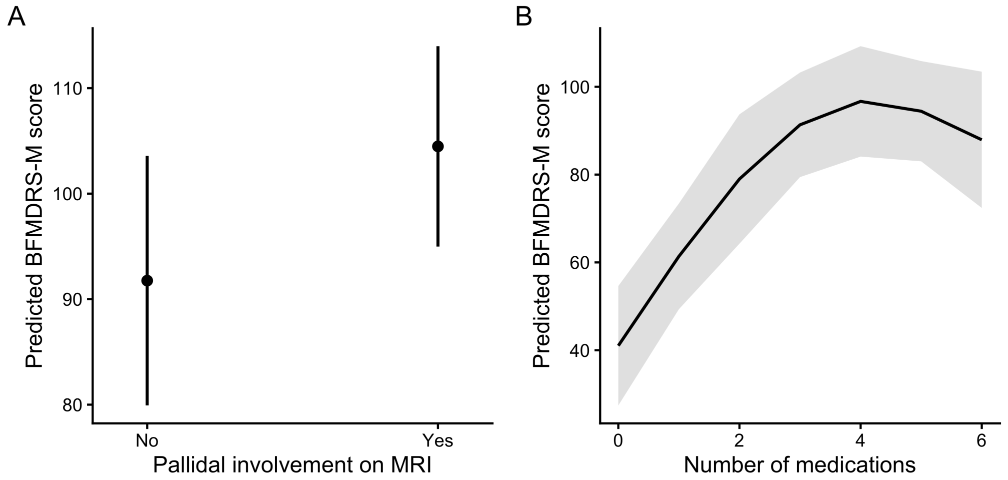

**Supplementary Figure 3. Model-predicted association between BFMDRS-M scores and pallidopallidal coherence in delta, alpha, beta and gamma.** Solid lines represent model-predicted BFMDRS-M scores from the adjusted mixed-effects models, with shaded areas indicating 95% confidence intervals. Points represent raw, unadjusted individual observations contributing to the model estimates. Colours indicate the number of medications.

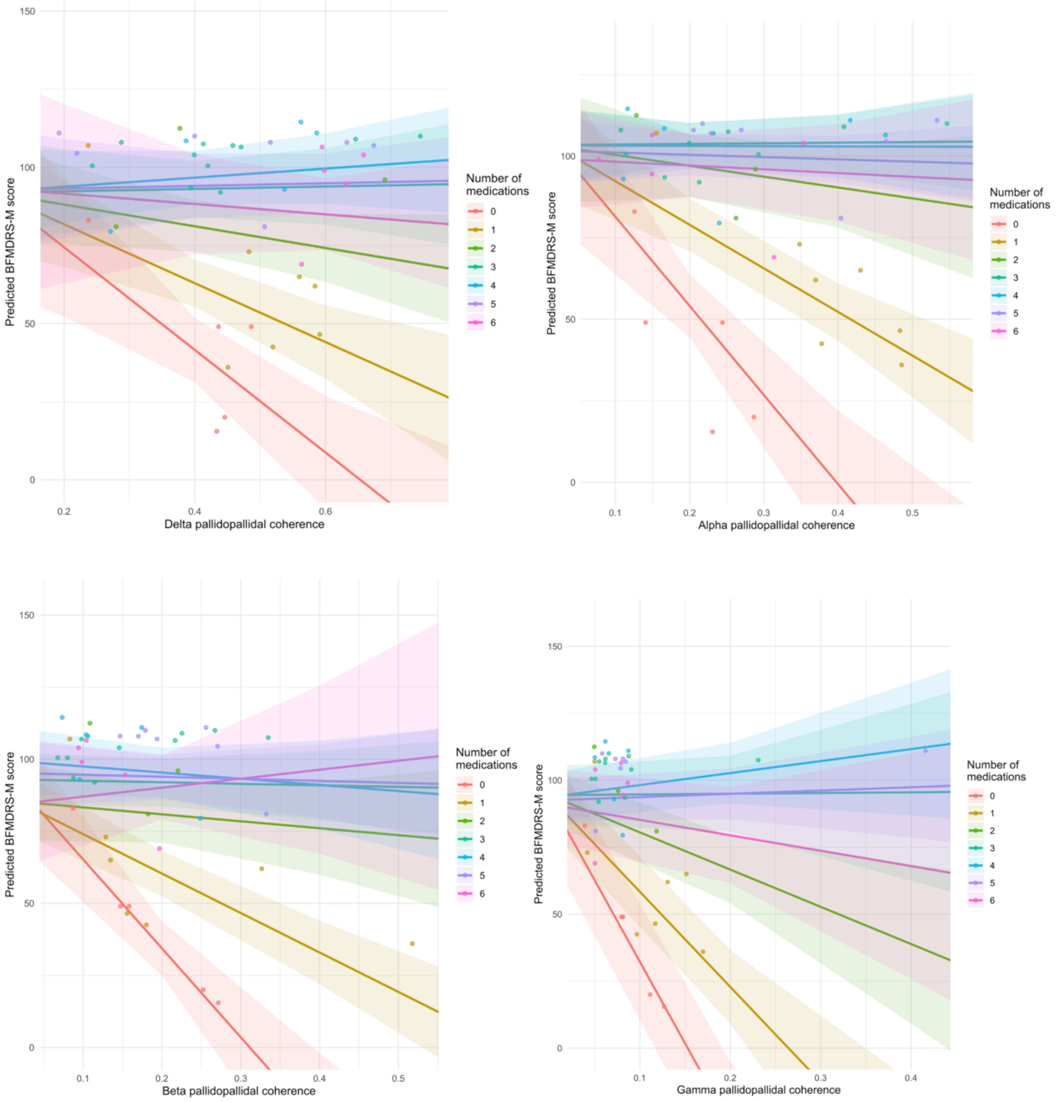

### Supplementary Section 4. Results for dystonia aetiology: Firth's Bias Reduced Logistic Regression

#### 4.1 Predictor: Peak power (extracted from best LFP channel)

##### 4.1.1 Unadjusted and adjusted models

**Supplementary Table 28. Results of Firth's bias-reduced logistic regression analyses on dystonia aetiology for other frequency bands.** Predictor: peak power (best LFP channel).

| Predictor | β | *SE* | OR | 95% CI for OR (lower) | 95% CI for OR (upper) | *P* value (uncorr.) | *P* value (FDR corrected) |
|---|---|---|---|---|---|---|---|
| **Unadjusted** | | | | | | | |
| Delta | -0.65 | 0.38 | 0.52 | 0.22 | 1.06 | 0.070 | 0.175 |
| Alpha | 1.41 | 2.31 | 4.10 | 0.06 | 1466.64 | 0.531 | 0.663 |
| Beta | -0.36 | 4.31 | 0.69 | 0.00 | 5665.54 | 0.933 | 0.933 |
| Gamma[b] | - | - | - | - | - | <0.001[b] | <0.001[b] |
| **Adjusted[a]** | | | | | | | |
| Delta | -0.58 | 0.46 | 0.55 | 0.17 | 1.39 | 0.215 | 0.652 |
| Alpha | 0.48 | 3.40 | 1.62 | 0.002 | 15821.60 | 0.900 | 0.900 |
| Beta | -3.37 | 5.99 | 0.03 | 0.00 | 24900.00 | 0.599 | 0.798 |
| Gamma[b] | - | - | - | - | - | - | - |

[a]Adjusted for pallidal involvement and medication number.
[b]Mixed-models results for gamma are not reported due to quasi-complete separation (see main manuscript). Results of the Fisher's exact test (with gamma peak power was binarised at a threshold of 0.01) are reported as the unadjusted model instead.
SE = standard error; OR = Odd Ratios; CI = confidence intervals; df = degrees of freedom; uncorr. = uncorrected; FDR = false discovery rate

#### 4.2 Predictor: Pallidopallidal coherence (extracted from best LFP channel)

##### 4.2.1 Unadjusted and adjusted models

**Supplementary Table 29. Results of Firth's bias-reduced logistic regression analyses on dystonia aetiology for other frequency bands.** Predictor: pallidopallidal coherence (best LFP channel).

| Predictor | β | *SE* | OR | 95% CI for OR (lower) | 95% CI for OR (upper) | *P* value (uncorr.) | *P* value (FDR corrected) |
|---|---|---|---|---|---|---|---|
| **Unadjusted** | | | | | | | |
| Delta | 3.17 | 2.99 | 23.98 | 0.07 | 13707.98 | 0.280 | 0.481 |
| Alpha | -1.27 | 3.30 | 0.27 | 0.00 | 210.99 | 0.700 | 0.700 |
| Beta | -5.31 | 4.80 | 0.004 | 0.00 | 53.13 | 0.258 | 0.481 |
| Gamma | -9.11 | 8.82 | 0.00 | 0.00 | 3088.53 | 0.289 | 0.481 |
| **Adjusted[a]** | | | | | | | |
| Delta | -1.49 | 4.24 | 0.22 | 0.00 | 1811.11 | 0.740 | 0.740 |
| Alpha | -4.92 | 4.83 | 0.01 | 0.00 | 100.89 | 0.322 | 0.628 |
| Beta | -4.79 | 5.43 | 0.008 | 0.00 | 308.83 | 0.377 | 0.628 |
| Gamma | -13.66 | 9.97 | 0.00 | 0.00 | 25855.62 | 0.216 | 0.628 |

[a]Adjusted for pallidal involvement and medication number.
SE = standard error; OR = Odd Ratios; CI = confidence intervals; df = degrees of freedom; uncorr. = uncorrected; FDR = false discovery rate

#### 4.3 Control analyses. Predictor: Peak power (best LFP channel) of 3 Hz narrow band around the peak

##### 4.3.1 Adjusted models

**Supplementary Table 30. Results of Firth's bias-reduced logistic regression analyses on dystonia aetiology for other frequency bands.** Predictor: peak power (best LFP channel).

| Predictor | β | *SE* | OR | 95% CI for OR (lower) | 95% CI for OR (upper) | *P* value (uncorr.) | *P* value (FDR corrected) |
|---|---|---|---|---|---|---|---|
| **Adjusted[a]** | | | | | | | |

| | | | | | | | |
|---|---|---|---|---|---|---|---|
| Delta | -0.99 | 0.75 | 0.37 | 0.05 | 1.66 | 0.195 | 0.487 |
| Theta | -0.63 | 0.73 | 0.52 | 0.01 | 1.75 | 0.363 | 0.605 |
| Alpha | 0.52 | 4.58 | 1.68 | 0.00 | 0.00 | 0.922 | 0.922 |
| Beta | -4.20 | 7.42 | 0.01 | 0.00 | 0.00 | 0.59 | 0.737 |
| Gamma[b] | - | - | - | - | - | <0.001 | <0.001 |

[a]Adjusted for pallidal involvement and medication number.
[b]Mixed-models results for gamma are not reported due to quasi-complete separation. Results of the Fisher's exact test (with gamma peak power was binarised at a threshold of 0.0095) are reported as the adjusted model instead (please note these are however not adjusted for any covariates).
SE = standard error; OR = Odd Ratios; CI = confidence intervals; df = degrees of freedom; uncorr. = uncorrected; FDR = false discovery rate

### 4.4 Control analyses. Predictor: Peak power (average across all LFP channels)

#### 4.4.1 Adjusted models

**Supplementary Table 31. Results of Firth's bias-reduced logistic regression analyses on dystonia aetiology for other frequency bands.** Predictor: peak power (best LFP channel).

| Predictor | β | *SE* | OR | 95% CI for OR (lower) | 95% CI for OR (upper) | *P* value (uncorr.) | *P* value (FDR corrected) |
|---|---|---|---|---|---|---|---|
| **Adjusted[a]** | | | | | | | |
| Delta | -0.52 | 0.43 | 0.59 | 0.22 | 1.54 | 0.258 | 0.430 |
| Theta | -1.55 | 1.05 | 0.21 | 0.01 | 1.96 | 0.169 | 0.422 |
| Alpha | -1.05 | 4.52 | 0.34 | 0.00 | 0.00 | 0.820 | 0.820 |
| Beta | -1.59 | 1.56 | 0.20 | 0.01 | 81.14 | 0.364 | 0.455 |
| Gamma[b] | - | - | - | - | - | <0.001 | <0.001 |

[a]Adjusted for pallidal involvement and medication number.
[b]Mixed-models results for gamma are not reported due to quasi-complete separation. Results of the Fisher's exact test (with gamma peak power was binarised at a threshold of 0.008) are reported as the adjusted model instead (please note these are however not adjusted for any covariates).
SE = standard error; OR = Odd Ratios; CI = confidence intervals; df = degrees of freedom; uncorr. = uncorrected; FDR = false discovery rate

### 4.5 Control analyses. Predictor: Pallidopallidal coherence (average across all LFP channels)

#### 4.5.1 Adjusted models

**Supplementary Table 32. Results of Firth's bias-reduced logistic regression analyses on dystonia aetiology for other frequency bands.** Predictor: peak power (best LFP channel).

| Predictor | β | *SE* | OR | 95% CI for OR (lower) | 95% CI for OR (upper) | *P* value (uncorr.) | *P* value (FDR corrected) |
|---|---|---|---|---|---|---|---|
| **Adjusted[a]** | | | | | | | |
| Delta | -1.17 | 6.17 | 0.30 | 0.00 | 1.57 | 0.860 | 0.860 |
| Theta | -4.12 | 6.13 | 0.01 | 0.00 | 14675.31 | 0.530 | 0.662 |
| Alpha | -5.76 | 6.59 | 0.01 | 0.00 | 4303.79 | 0.411 | 0.662 |
| Beta | -8.83 | 8.92 | 0.00 | 0.00 | 1966.73 | 0.308 | 0.662 |
| Gamma | -22.49 | 16.35 | 0.00 | 0.00 | 0.00 | 0.221 | 0.662 |

[a]Adjusted for pallidal involvement and medication number.
SE = standard error; OR = Odd Ratios; CI = confidence intervals; df = degrees of freedom; uncorr. = uncorrected; FDR = false discovery rate

#### 4.6 Figures

**Supplementary Figure 4. Model-predicted association between dystonia aetiology and A) pallidal involvement and B) medication burden.** Graphs are based on the model using theta peak power (best LFP channel) as a predictor, but the estimated shape of the association is consistent across all models.

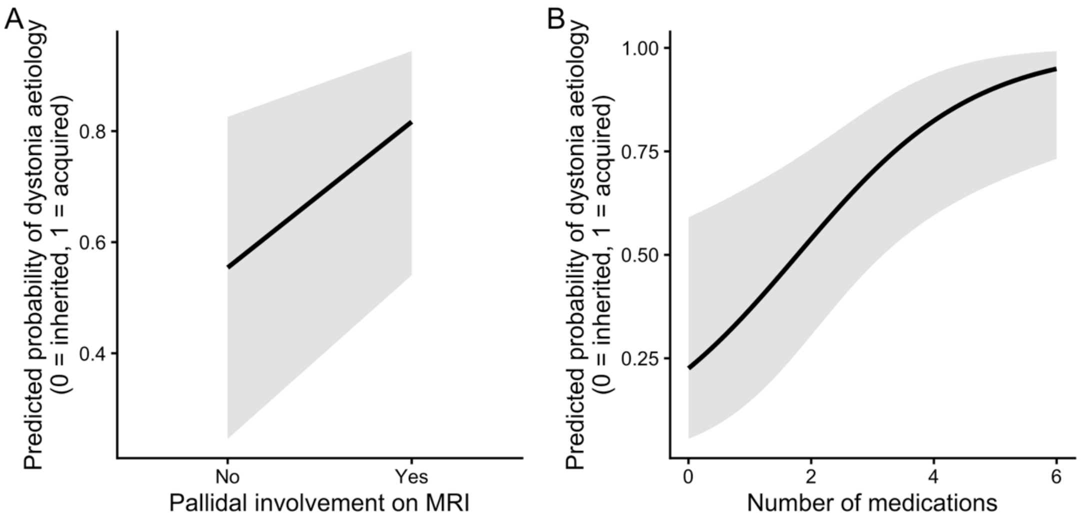
