## Supplementary Material - Brainsense_ecg_removal method description and synthetic validation for "Pallidal Local Field Potentials markers of dystonia aetiology and severity in children"

### BRAINSENSE_ECG_REMOVAL FUNCTION

Automated cardiac artefact template identification and subtraction from BrainSense LFP data, based on Hammer et al. (2022).

**Author:** Irene Faiman

#### 1. Template Seed Identification

To identify an initial cardiac artefact template seed, the first 5 seconds of the recording were treated as a search window. Within this, all possible contiguous epochs of fixed duration (default: 750 ms) were extracted using a sliding window with one-sample increments. The epoch length was chosen based on normative paediatric scores (<16 years; Dickinson, 2005) to accommodate the upper physiological bounds of the PR and QT intervals (200ms and 450ms respectively), plus a 100ms buffer.

Each candidate epoch (template) was mean-centred and normalised by its Euclidean ($L^2$) norm to remove baseline offsets and amplitude differences. This ensured that subsequent comparison reflected waveform shape rather than signal energy.

Each candidate template was then time-reversed and used in a matched filter (conv() function) against the full LFP signal. Peaks in the matched filter output with a minimum RR interval of 0.5 seconds (corresponding to a maximum heart rate of 120 bpm; Ostchega et al., 2011) were identified using the Matlab findpeaks() function. The candidate template with the highest average matched-filter peak amplitude was selected as a preliminary cardiac template seed, under the assumption that true cardiac artefacts recur periodically and produce strong, repeated correlations. The preliminary template was then used to detect preliminary cardiac artefact locations in the full signal by re-running the convolution and findpeaks() functions; the detected locations were time-aligned, and the initial cardiac template seed was computed as the average of all locations where the artefact was detected.

**Fig. 1. Template seed identification procedure.** For a channel where cardiac artefact is present (left) and one where it is not (right), subplots depict (top) the best single normalised epoch selected as the preliminary template seed; (centre) the matched-filter output (red) applied to the raw LFP (black) with detected artefact peak locations (black dots); (bottom) overlaid detected preliminary cardiac artefact epochs (grey) with the resulting averaged template seed (blue).

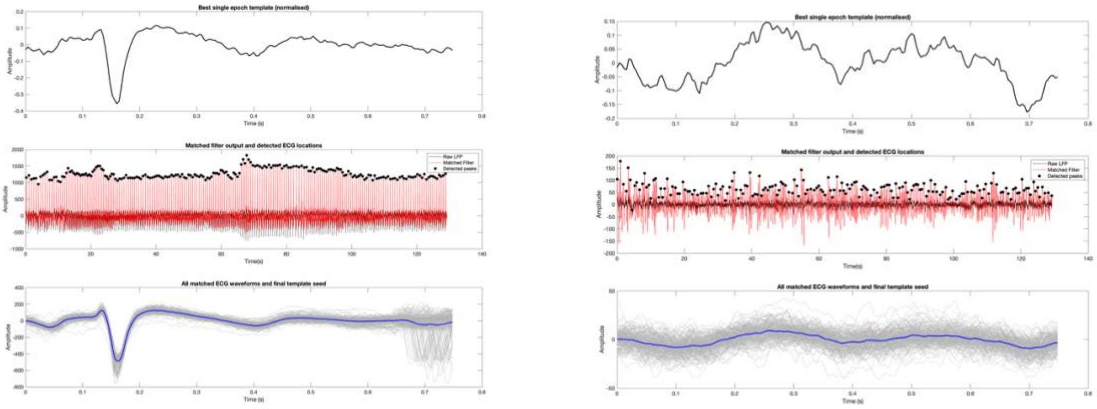

### 2. Modified Woody's Adaptive Filter

An iterative refinement procedure consistent with a Modified Woody’s filter was applied to the initial template seed to further improve cardiac artefact localisation and template estimation. At each iteration, the template estimate was normalised, time-reversed, and convolved with the full LFP signal to generate a matched filter output.

Candidate artefact occurrences were identified as local maxima exceeding the 97.5$^{th}$ percentile of the matched filter output and separated by at least 0.5 seconds. The detected artefact locations were time-aligned and averaged to form an updated template. This process was repeated until convergence was reached.

To assess convergence, the average cosine similarity (i.e., normalised dot product of mean-centred signals) between the current template and each artefactual epoch was computed, in line with similar Woody implementations (Silva, 2026). Convergence was defined as a positive improvement in average cosine similarity that fell below the set tolerance level of 0.005. If the improvement became negative, indicating deterioration of the template, the algorithm terminated and retained the previous template estimate.

**Fig 2. Artefactual locations detected following Modified Woody's Adaptive Filter.** For a channel where cardiac artefact is present (top) and one where it is not (bottom), raw LFP signal (black) with artefact locations identified following iterative template refinement (Modified Woody’s filter).

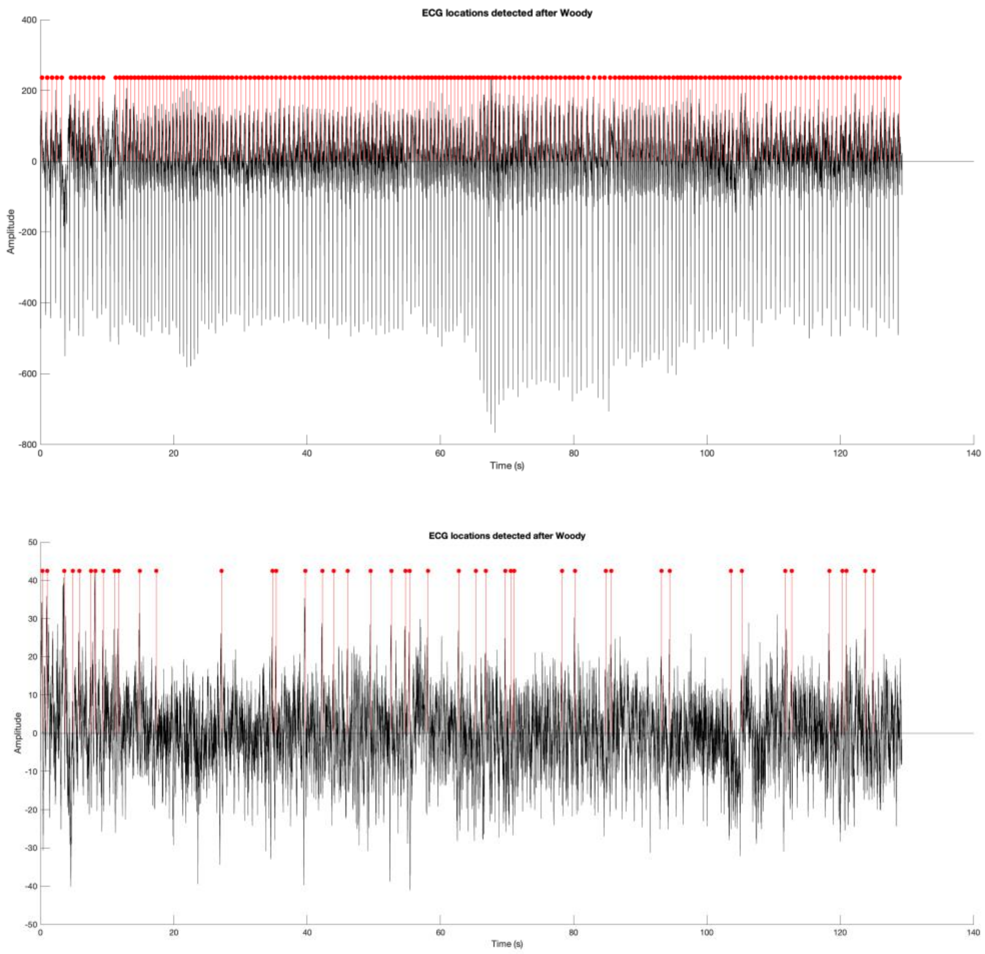

#### 3. Visual template review including physiological plausibility check

The resulting detected peaks were subjected to a physiological plausibility check by estimating heart rate (beats per minute, BPM) from the number of detected R-peaks. The BPM estimate was compared against liberal age-based physiological bounds (48–187 BPM) derived from paediatric data (Riopel et al., 1979; Ostchega et al., 2021), and warnings were generated if the estimated heart rate fell outside these limits.

For qualitative validation, all detected artefactual epochs were plotted alongside the final Woody-adapted template. Following visual inspection, the user manually confirmed whether the detected events resembled true cardiac activity. If approved, cardiac artefact correction proceeded through the following steps. If rejected, the correction step was aborted and the original (uncorrected) LFP signal was retained for further analysis.

**Fig 3. Visual template review step.** For a channel where cardiac artefact is present (left) and one where it is not (right), all detected artefactual instances (grey) plotted alongside the final Woody-adapted template (blue). The estimated BPM derived from detected peaks is displayed. The user is asked to assess whether the template resembles a cardiac artefact.

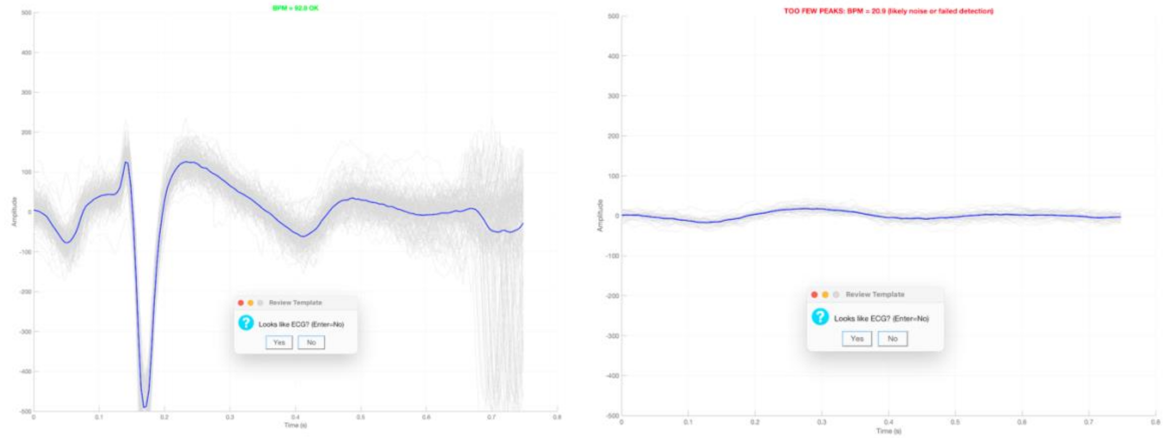


#### 4. Forced Search for Missed Artefacts Identification

To identify potentially missed cardiac artefacts, RR intervals were computed from peak locations, and intervals exceeding 1.5 times the modal RR interval were considered indicative of potential missed detections.

For each identified gap, the number of potentially missing artefacts was estimated.

The strongest correlation peaks within the gap were identified using matched filter and peak-finding functions, constrained by the estimated number of missing beats and a minimum peak distance corresponding to the physiological RR interval.

**Fig 4. Additional artefact locations detected through forced search.** For files with user-confirmed cardiac activity, raw LFP signal (black) with original Woody detections (red) and additional peaks identified through gap-based forced matched-filter search (blue), highlighting artefacts recovered from abnormally long RR intervals.

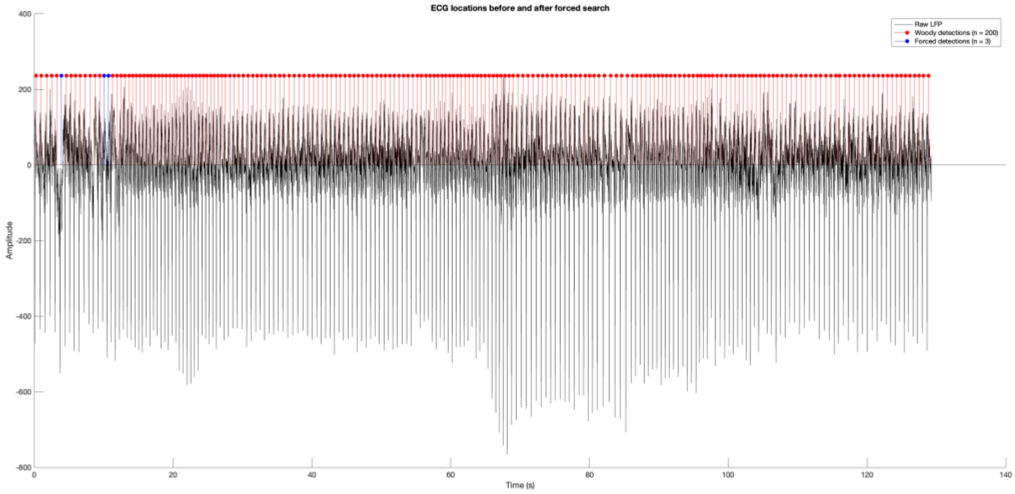


### 5. Subtraction of artefact template

Cardiac artefacts were removed from the LFP by subtracting the final Woody-adapted template at each detected artefact location. Boundary checks were applied to trim the template if the window extended beyond signal limits. To prevent double subtraction in cases of overlapping template windows, these were adaptively trimmed at the edges.

**Fig 5. Subtraction of artefact template**. Top left: Raw (black) and cleaned (red) LFP signals following template subtraction. Bottom-left: Difference (subtracted) signal illustrating removed cardiac components. Right: Power spectral density of raw (black) and cleaned (blue) signals (dB/Hz), depicting spectral changes following artefact correction.

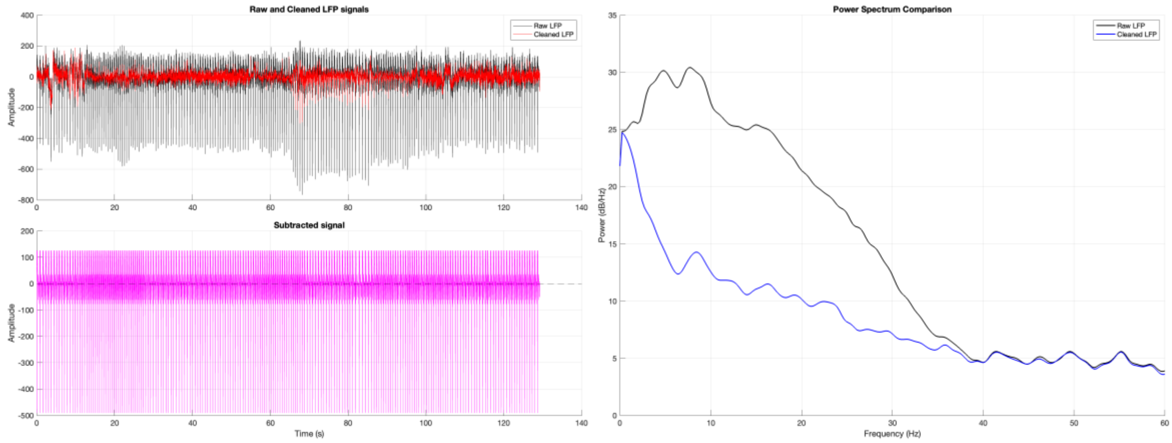

**BRAINSENSE_ECG_REMOVAL FUNCTION VALIDATION**

**Assessment of performance with synthetic cardiac artefact superimposed to real, non-artefactual LFP signal**

**Methods:**

To assess the performance of the brainsense_ecg_removal function, synthetic cardiac artefacts were added to a real, single-channel LFP recording that was known not to contain cardiac artefacts. To simulate physiologically plausible heartbeats, RR intervals were drawn from a Gaussian distribution with mean 80 bpm, and SD 70 ms. A QRS template was inserted at each simulated beat (amplitude 200 µV), producing a contaminated LFP signal (Fig 6 and 7). This was processed with the brainsense_ecg_removal function (Fig 8 and 9). Performance was assessed in the following ways:

1. by correlating the original LFP signal to the final cleaned LFP both throughout the whole signal and at cardiac artefact locations only
2. by computing the Root Mean Square Error (RMSE) between the original LFP signal and the final cleaned signal (following brainsense_ecg_removal pipeline). RMSE provides a measure of the average amplitude difference between signals, therefore quantifying the magnitude of error introduced by the artefact removal procedure. We separately estimated RMSE throughout the whole signal, at cardiac artefact locations only, and at non-artefact regions only.

**Fig 6. Synthetic cardiac artefact template**

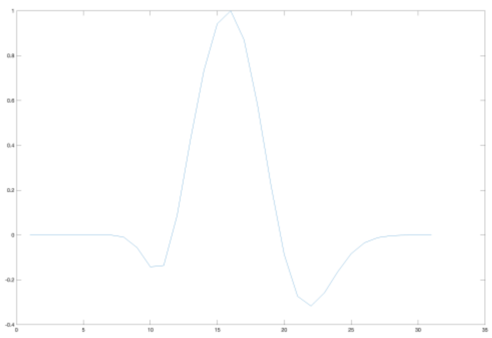

**Fig 7. Synthetic cardiac artefact added to real LFP data**. Displayed are the first five seconds of recordings.

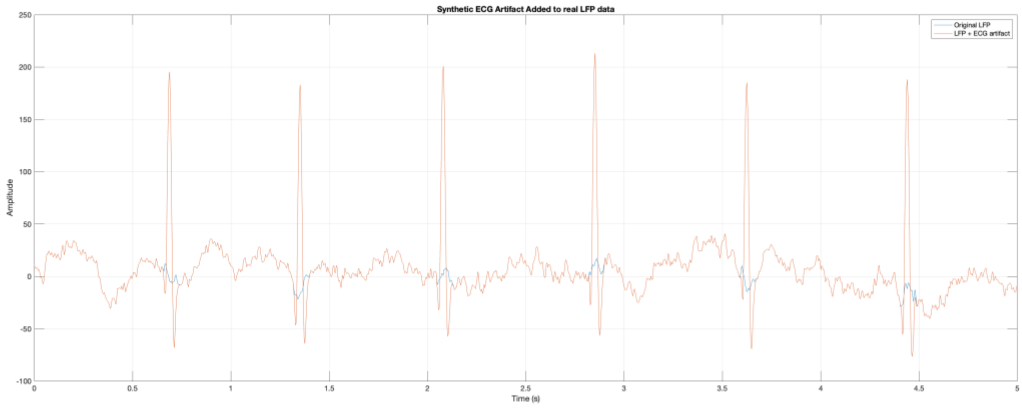

**Fig 8. Visual review step of brainsense_ecg_removal function.** Displayed is the final template generated by the function prior to subtraction.

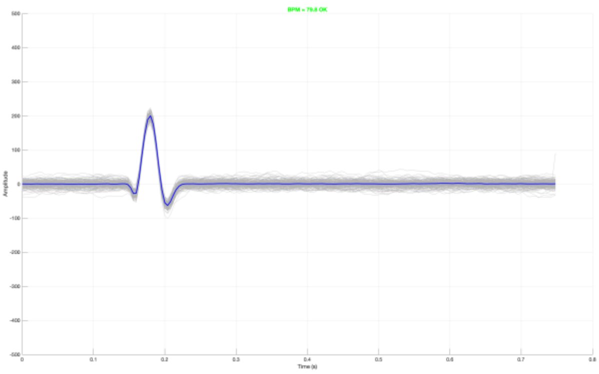


**Fig 9. Output figure of brainsense_ecg_removal function.** Displayed are the raw and cleaned LFP tracks (top left), the subtracted signal (bottom left), and the power spectrum difference between the raw and cleaned LFP signals (right).

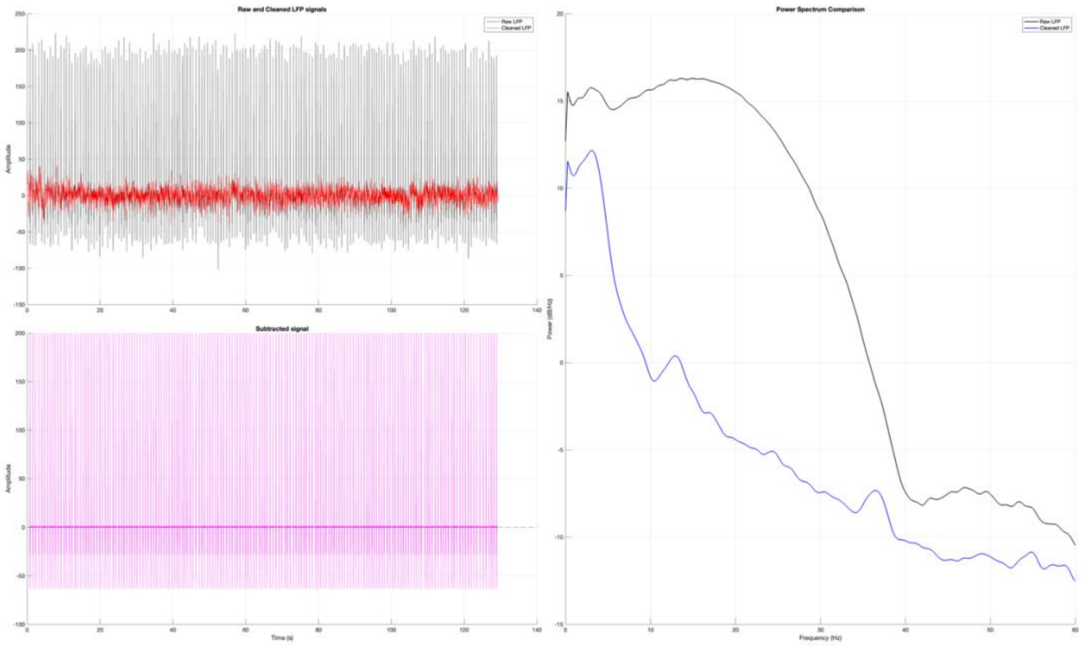


### Results:

The cardiac artefact removal algorithm effectively suppressed the simulated artefacts while preserving the underlying LFP signal (Fig 10 and 11). The cleaned signal showed high correlation with the original LFP signal both globally (r = 0.9987) and when considering ECG artefact locations only (r = 0.9993). RMSE was minimal overall (0.57 µV), at cardiac artefact locations (0.38 µV), and in non-artefact regions (0.60 µV), indicating successful artefact suppression with negligible impact on the underlying neural signal.

**Fig 10 Original, ECG-contaminated, and cleaned LFP signals for a single heartbeat.** Y axis = amplitude.

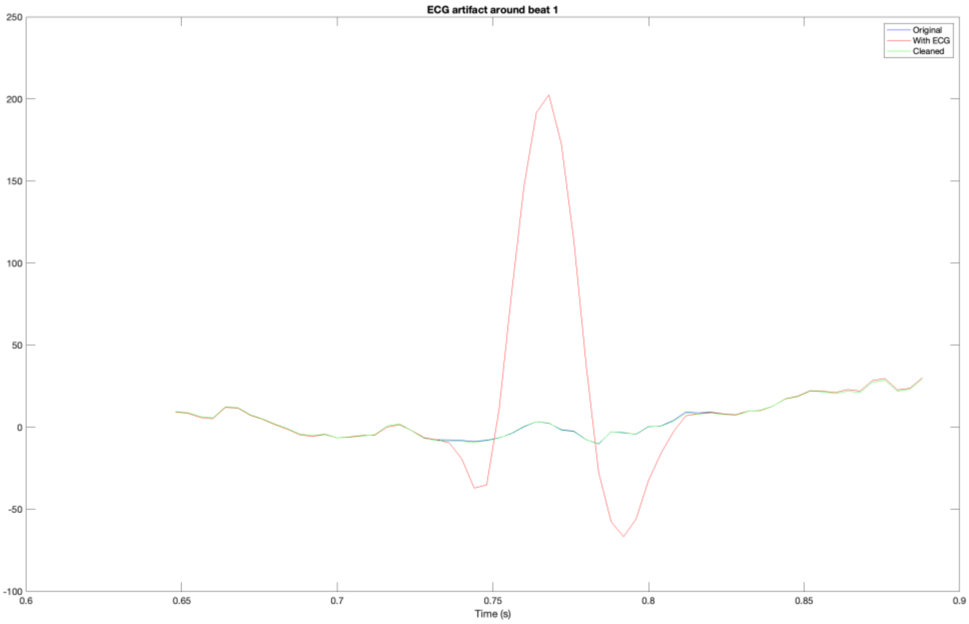


**Fig 11 Original, ECG-contaminated, and cleaned LFP signals for the first five heartbeats in the recordings.** Y axis = amplitude.

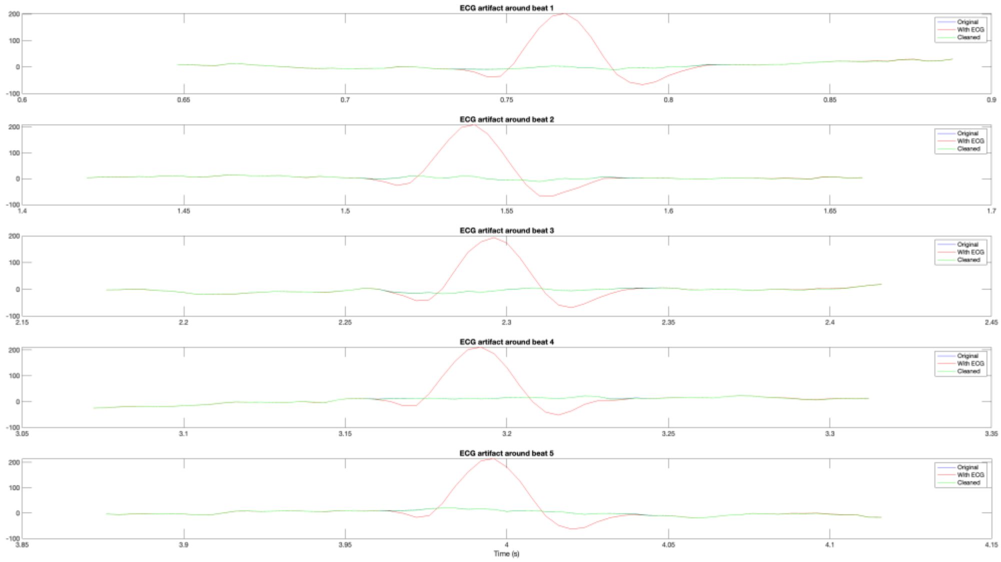
